# The Impact on Clinicians of Recent Changes to the U.S. Immunization Schedule

**DOI:** 10.64898/2026.11.20.26368496

**Authors:** Matthew Z. Dudley, William J. Moss, Joshua M. Sharfstein, Michelle Goryn, Rose Weeks, Hannah Melchinger, Daniel A. Salmon

## Abstract

**Importance:** While recent changes in federal vaccine policy may have negatively impacted clinician vaccination efforts, data assessing such impacts are limited.

**Objective:** To evaluate the impact on clinicians of these changes to vaccine recommendations.

**Design:** A high-quality panel survey was administered in July 2026 and compared to a similar survey administered in January 2023.

**Setting:** The panel consisted of clinicians from across the United States.

**Participants:** Respondents included 508 pediatricians, 500 family physicians, 166 nurse practitioners (NPs), 167 physician assistants (PAs), and 166 nurses.

**Main Outcomes and Measures:** Primary outcomes of interest included clinician strength of vaccine recommendations for patients and trusted sources of vaccine information and recommendations.

**Results:** Most clinicians trusted vaccine recommendations from professional medical organizations such as the American Academy of Pediatrics (AAP) (91%) and the American Academy of Family Physicians (AAFP) (90%), whereas fewer than half of clinicians trusted vaccine recommendations from federal agencies such as the Centers for Disease Control and Prevention (CDC) (47%) or the Food and Drug Administration (FDA) (42%). Compared to 2023, far fewer clinicians reported trusting vaccine information from CDC (47% vs 79%) or FDA (43% vs 75%) (adjusted p<0.01).

Despite reduced trust in federal vaccine recommendations, most clinicians continued to recommend routine vaccines for their patients, with the notable exception of COVID-19, for which far fewer clinicians recommend vaccination compared to 2023 regardless of patient age, but especially among 12–15-year-olds (41% vs 71%, p<0.01) and 16–24-year-olds (43% vs 74%, p<0.01). Many clinicians also reported noticing substantial increases in the frequency (80%) and intensity (76%) of patient vaccine concerns over the past year, as well as in their time spent discussing vaccines with patients (61%) and looking up vaccine information (45%).

**Conclusions and Relevance:** This survey provides insight into the point-of-care impact of recent changes in federal vaccine policy and the shifts in clinician vaccine recommendations and trusted sources of vaccine information since the end of the pandemic. Professional associations can fill the current void in federal leadership by providing clinicians with reliable vaccine information and recommendations.

**Key Points:** *Question:* Have the recent changes in federal vaccine policy impacted clinician vaccination efforts and patient encounters?

*Findings:* This large, high-quality panel survey found that many clinicians have turned to nongovernmental sources of vaccine information and recommendations as their trust in federal agencies has diminished. Despite this, most clinicians continue to recommend routine vaccines for their patients, with the notable exception of COVID-19. Clinicians also reported increases in patient vaccine hesitancy and time spent counseling patients about vaccines.

*Meaning:* Most clinicians continue to support vaccination despite fluctuations in federal vaccine recommendations, largely relying on professional medical organizations for vaccine guidance.

## Introduction

Vaccine recommendations made by the Advisory Committee on Immunization Practices (ACIP) to the U.S. Centers for Disease Control and Prevention (CDC) were historically endorsed by professional medical organizations, delivering one set of harmonized recommendations. However, this traditional cohesion was disrupted following the unconventional overhaul of ACIP by the Department of Health and Human Services (HHS) in June 2025.^1,2^ Over the next six months, the new ACIP altered recommendations for several vaccines, including hepatitis B, influenza, measles-mumps-rubella-varicella (MMRV), and COVID-19.^3^ Then, in January 2026, the CDC’s immunization schedule was updated based on a presidential memorandum without a corresponding ACIP recommendation, redesignating certain vaccines (e.g., hepatitis A, hepatitis B, influenza, meningococcal, rotavirus) as no longer routinely recommended but instead available based on shared clinical decision-making (SCDM).^4^ These changes were criticized by public health experts,^5–9^ prompting prominent medical organizations such as the American Academy of Pediatrics (AAP) to recommit to providing their own vaccine recommendations independently of CDC,^10^ with many states choosing to follow AAP’s recommendations instead of CDC’s.^11^ In March 2026, a U.S. district court blocked these recent changes to CDC’s vaccine recommendations,^12^ though this ruling was appealed by HHS.

Turmoil in vaccine policy may confuse clinicians,^13,14^ and since clinician recommendations strongly correlate with vaccination,^15,16^ declines in clinician vaccine confidence may have downstream effects on vaccine coverage. Misleading public statements from federal leadership may have also undermined public vaccine confidence and coverage.^17^ However, data assessing the impact of these events on clinician vaccine recommendations have been limited.

We report the results of a high-quality panel survey of clinicians evaluating how recent events have affected their vaccination efforts. We also examine changes in clinician responses since our last survey, conducted near the end of the COVID-19 pandemic.

## Methods

### Recruitment

This survey was administered between June 24 and July 30, 2026 to clinicians recruited by Konovo Healthcare Intelligence, a leading healthcare research firm.^18^ Konovo offers uniquely high-quality data for U.S. clinicians via a double opt-in network panel that samples from a population of more than 2 million clinicians regularly updated and validated from standard core sources.

Recruitment aimed to achieve a total sample of 1,500 primary care clinicians who administered vaccines. In addition to pediatricians and family physicians, eligible clinicians included nurse practitioners (NPs), physician assistants (PAs), and nurses, provided they specialized in pediatrics, family practice, or internal medicine. Of the 13,990 such panel members who were invited, 3,032 agreed to participate, corresponding to a response rate of 22% (Supplemental Table 1). Responders and non-responders differed in their clinician type, region, and gender (Supplemental Table 2). Quotas were set for three groups (pediatricians, family physicians, and NPs/PAs/nurses) of roughly equal size (about 500 each), with at least 300 clinicians from each of the four major U.S. regions (Midwest, Northeast, South, West), to ensure sufficient power for comparisons. Clinicians were incrementally administered the survey until all quotas were met.

Participants consented to the survey, which was voluntary and anonymous. The Johns Hopkins Bloomberg School of Public Health Institutional Review Board (IRB) considered this study public health surveillance and not human subjects research.

### Survey Content

The survey instrument (Supplemental Material 1) replicated relevant items from a prior survey conducted using the same panel in January 2023 to facilitate comparisons over time,^19^ while also including new items pertinent to the current environment. Repeat survey items assessed patient and practice characteristics, strength of vaccine recommendations for patients, prevalence of patient vaccine concerns, preparedness to discuss vaccines with patients, utility of vaccine resources, and trusted sources of vaccine information. Both surveys assessed trust in CDC using a validated 14-item scale.^20^ Both surveys assessed familiarity with and use of the Vaccine Adverse Event Reporting System (VAERS)^21^ among half of respondents, with the other half asked about the non-existent, fabricated Immunization Adverse Reaction Monitoring (IARM) system to measure social desirability bias.

New survey items assessed trust in vaccine recommendations from various sources, time spent researching vaccine information and discussing vaccines with patients, and patient vaccine uptake and hesitancy. Items were added to assess clinician vaccine recommendations for patients with more specificity than the previous survey (which largely focused on COVID-19). To allow this while maintaining comparability with our previous survey, half of respondents were randomized to receive the new (more specific) survey items while the other half received the old (more general) items. Respondents were asked whether they provided care for children, young adults, and/or older adults, and were only presented survey items pertaining to vaccines relevant for their patient populations. Most items used 4- or 5-point scales as response options to improve precision.

### Data Analysis

Survey responses were summarized and stratified by trust in CDC vaccine recommendations. Changes in response frequency between surveys were calculated for repeat items. Respondents from the 2023 survey who would not have been eligible for the current survey (e.g., pharmacists) were not included in the comparative analysis to ensure the sampling strategy between the two survey waves was aligned, with both samples consisting of pediatricians, family physicians, and NPs/PAs/nurses split into three evenly numbered strata. Differences between stratifications and timepoints were assessed using the Kruskal-Wallis test for skewed continuous variables and the chi-square test for categorical and binary variables (p<0.05 considered statistically significant). Both crude and adjusted differences between timepoints, along with corresponding 95% confidence intervals (CIs), were calculated to control for potential confounding of clinician characteristics shown to differ significantly between timepoints after using backwards stepwise regression to reduce collinearity (i.e., specialty, region, age range of patient population, years since graduation) (Supplemental Table 3).

A composite, linear score was generated for the general trust in CDC construct scale,^20^ scoring the 4-point Likert scale response options 0-3 (and reversing the score for negatively worded items). The scale score numerator summed the scored responses, with the denominator being the total possible score (accounting for missing data), creating a scale from 0 (complete lack of trust) to 100 (complete trust). A binary version of this scale score dichotomized at the midpoint (50).

Data were analyzed using Stata (version 16).^22^

## Results

### Survey Sample

Characteristics of the 1,507 clinicians surveyed are outlined in Table 1, with additional details on their patient populations in Supplemental Table 4. About half (51%) of participating clinicians were in private practice, with 21% in a hospital or medical center, 15% in a practice network or health maintenance organization, and 11% in a community health center or federally qualified health center. About half (48%) practiced in a suburban setting, with 36% in an urban setting and 16% in a rural setting.

**Table 1.** Characteristics of Participating Clinicians, their Practices, and their Patient Populations, Stratified by Trust in CDC Vaccine Recommendations.

|  | Total<br>N (%)* | Trust in CDC Vaccine Recommendations |  | p-value |
|---|---|---|---|---|
|  |  | Low Trust<br>N (%)** | High Trust<br>N (%)** |  |
| All | N=1,507 | 801 (53.2%) | 704 (46.8%) |  |
| **Clinician Characteristics** |  |  |  |  |
| Clinician Type |  |  |  | **<0.01** |
| Pediatrician | 508 (33.7%) | 357 (70.3%) | 151 (29.7%) |  |
| Family Physician | 500 (33.2%) | 243 (48.7%) | 256 (51.3%) |  |
| Nurse Practitioner (NP) | 166 (11.0%) | 59 (35.5%) | 107 (64.5%) |  |
| Physician Assistant (PA) | 167 (11.1%) | 86 (51.5%) | 81 (48.5%) |  |
| Nurse | 166 (11.0%) | 56 (33.9%) | 109 (66.1%) |  |
| Specialty |  |  |  | **<0.01** |
| Pediatrics | 622 (41.3%) | 409 (65.8%) | 213 (34.2%) |  |
| Family Practice | 810 (53.7%) | 362 (44.8%) | 446 (55.2%) |  |
| Internal Medicine | 75 (5.0%) | 30 (40.0%) | 45 (60.0%) |  |
| Highest completed clinical degree |  |  |  | **<0.01** |
| LPN Certificate | 2 (0.1%) | 1 (50.0%) | 1 (50.0%) |  |
| Associate | 26 (1.7%) | 8 (30.8%) | 18 (69.2%) |  |
| Bachelor's | 108 (7.2%) | 36 (33.6%) | 71 (66.4%) |  |
| Master's | 319 (21.2%) | 143 (44.8%) | 176 (55.2%) |  |
| Doctoral | 1,051 (69.8%) | 613 (58.4%) | 437 (41.6%) |  |
| Years since graduation |  |  |  | **<0.01** |
| <10 | 451 (29.9%) | 215 (47.7%) | 236 (52.3%) |  |
| 10-19 | 446 (29.6%) | 258 (58.1%) | 186 (41.9%) |  |
| 20-29 | 330 (21.9%) | 187 (56.7%) | 143 (43.3%) |  |
| 30-39 | 196 (13.0%) | 106 (54.1%) | 90 (45.9%) |  |
| 40+ | 84 (5.6%) | 35 (41.7%) | 49 (58.3%) |  |
| Region |  |  |  | **<0.01** |
| Midwest | 349 (23.2%) | 182 (52.1%) | 167 (47.9%) |  |
| Northeast | 337 (22.4%) | 178 (52.8%) | 159 (47.2%) |  |
| South | 473 (31.4%) | 229 (48.5%) | 243 (51.5%) |  |
| West | 348 (23.1%) | 212 (61.1%) | 135 (38.9%) |  |
| Race/Ethnicity |  |  |  | **<0.01** |
| Asian | 208 (14.8%) | 110 (52.9%) | 98 (47.1%) |  |
| Black | 56 (4.0%) | 25 (44.6%) | 31 (55.4%) |  |
| Hispanic/Latino | 59 (4.2%) | 21 (35.6%) | 38 (64.4%) |  |
| White | 1,042 (73.9%) | 560 (53.8%) | 480 (46.2%) |  |
| Other/Multiple | 45 (3.2%) | 32 (71.1%) | 13 (28.9%) |  |
| Gender |  |  |  | 0.87 |
| Male | 599 (41.2%) | 318 (53.1%) | 281 (46.9%) |  |
| Female | 854 (58.8%) | 456 (53.5%) | 396 (46.5%) |  |
| Candidate supported in 2024 presidential election |  |  |  | **<0.01** |
| Donald Trump (Republican Party) | 265 (25.7%) | 103 (39.0%) | 161 (61.0%) |  |
| Kamala Harris (Democratic Party) | 607 (58.8%) | 408 (67.2%) | 199 (32.8%) |  |
| Other | 160 (15.5%) | 70 (43.8%) | 90 (56.3%) |  |
| **Practice Characteristics** |  |  |  |  |
| Practice Setting |  |  |  | 0.36 |
| Private Practice | 761 (50.5%) | 420 (55.3%) | 339 (44.7%) |  |
| Practice Network or Health Maintenance Organization (HMO) | 228 (15.1%) | 120 (52.6%) | 108 (47.4%) |  |
| Hospital or Medical Center | 319 (21.2%) | 156 (48.9%) | 163 (51.1%) |  |
| Community Health Center or Federally Qualified Health Center (FQHC) | 167 (11.1%) | 90 (53.9%) | 77 (46.1%) |  |
| Other | 32 (2.1%) | 15 (46.9%) | 17 (53.1%) |  |
| Urbanicity of Practice Location |  |  |  | 0.87 |
| Rural | 241 (16.0%) | 130 (53.9%) | 111 (46.1%) |  |
| Suburban | 724 (48.0%) | 388 (53.7%) | 335 (46.3%) |  |
| Urban | 542 (36.0%) | 283 (52.3%) | 258 (47.7%) |  |
| Average number of patients per day |  |  |  | **<0.01** |
| <10 | 46 (3.1%) | 19 (41.3%) | 27 (58.7%) |  |
| 10-24 | 962 (63.8%) | 547 (56.9%) | 414 (43.1%) |  |
| 25+ | 499 (33.1%) | 235 (47.2%) | 263 (52.8%) |  |
| dPractice uses Electronic Health Records (EHR) | 1,482 (98.3%) | 786 (98.1%) | 694 (98.6%) | 0.49 |
| Practice participates in the Vaccines for Children (VFC) program | 950 (63.0%) | 549 (68.5%) | 400 (56.8%) | **<0.01** |
| Seasonal Influenza Vaccination Available in Practice |  |  |  |  |
| 2023-2024 | 1,451 (96.3%) | 773 (96.5%) | 676 (96.0%) | 0.62 |
| 2024-2025 | 1,463 (97.1%) | 781 (97.5%) | 680 (96.6%) | 0.29 |
| 2025-2026 | 1,464 (97.1%) | 781 (97.5%) | 681 (96.7%) | 0.37 |

\* Column percentages out of total sample (N=1,507)
\*\*Row percentages among subsample with data on trust in CDC vaccine recommendations (N=1,505); N=2 survey respondents reported being unfamiliar with CDC vaccine recommendations and were thus counted as missing for this variable

### Trusted Sources of Vaccine Information

The sources of vaccine information most frequently used by clinicians include professional medical organization websites (71%) and online medical resources such as UpToDate and Medscape (62%) (Supplemental Figure 1). The sources of vaccine information most trusted by clinicians included UpToDate (89%), academic/medical journals (87%), professional medical organizations (86%), and academic/medical institutions (85%) (Figure 1). Fewer than half of clinicians trusted vaccine information from the CDC (47%) and FDA (43%).

**Figure 1.**
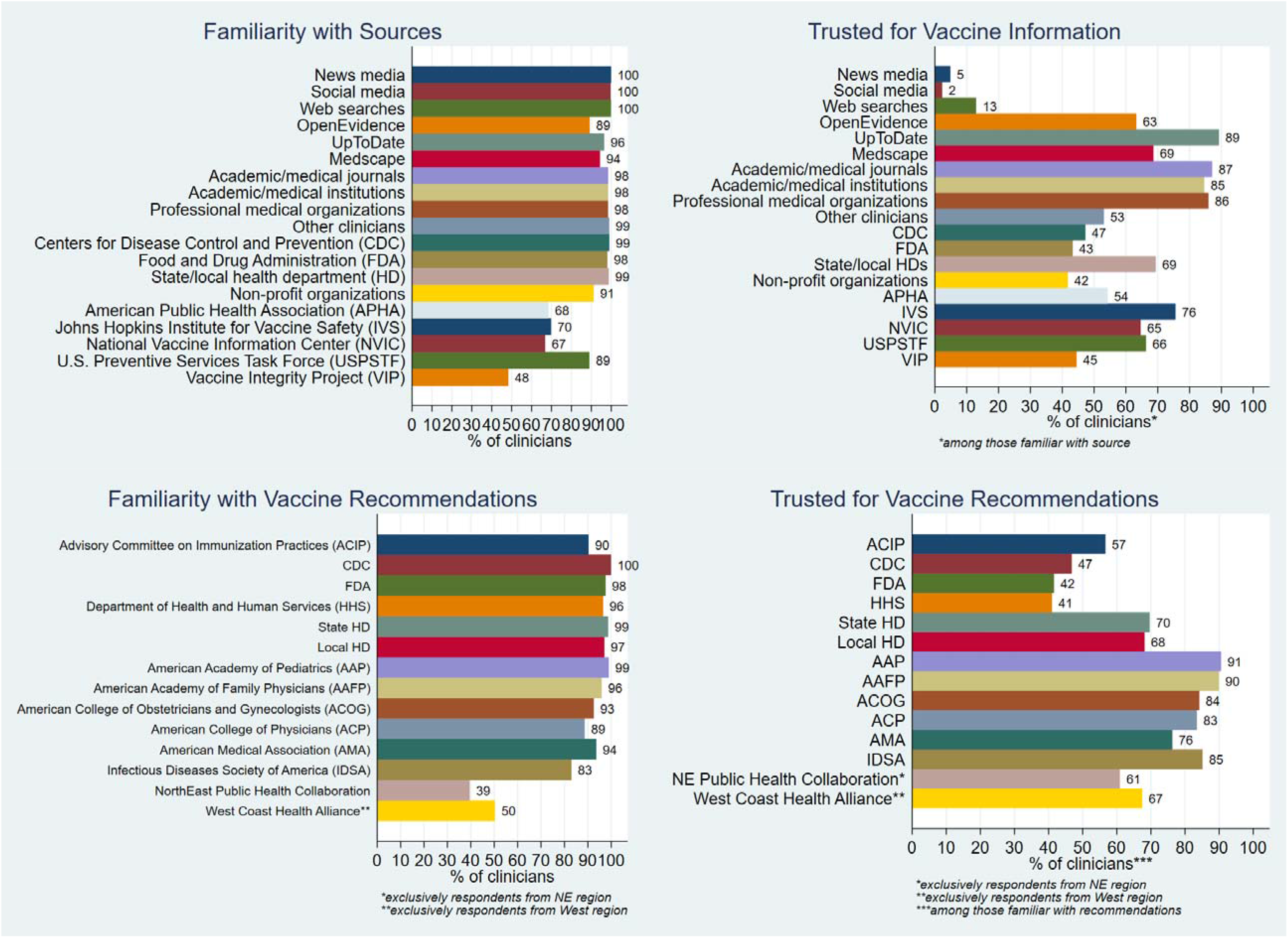
Clinician Familiarity with and Trust in Sources of Vaccine Information and Recommendations

### Trusted Sources of Vaccine Recommendations

The vaccine recommendations most trusted by clinicians were from professional medical organizations such as AAP (91%) (Figure 1). Fewer than half of clinicians trusted vaccine recommendations from federal sources such as CDC (47%), FDA (42%), and HHS (41%). At least half of clinicians were unfamiliar with regional state coalitions issuing vaccine recommendations, even among clinicians from those specific regions.

### Clinician Vaccine Recommendations for Patients

Most clinicians continued to recommend routine vaccines to their pediatric patients (Figure 2). Nearly all (at least 97%) recommended *Haemophilus influenzae* type b (Hib) vaccine, inactivated polio vaccine (IPV), meningococcal conjugate vaccine (MenACWY), pneumococcal conjugate vaccine (PCV), diphtheria/tetanus/acellular pertussis combination vaccine (DTaP/Tdap), measles/mumps/rubella (MMR) combination vaccine, and varicella vaccine. Other pediatric vaccines such as hepatitis A, hepatitis B, rotavirus, respiratory syncytial virus (RSV), and MMRV were recommended by 89-92% of clinicians. Strong pediatric vaccine recommendations were most common for MMR (96%), DTaP/Tdap (95%), Hib (94%), IPV (93%), PCV (93%), and varicella (93%) (Supplemental Table 5).

**Figure 2.**
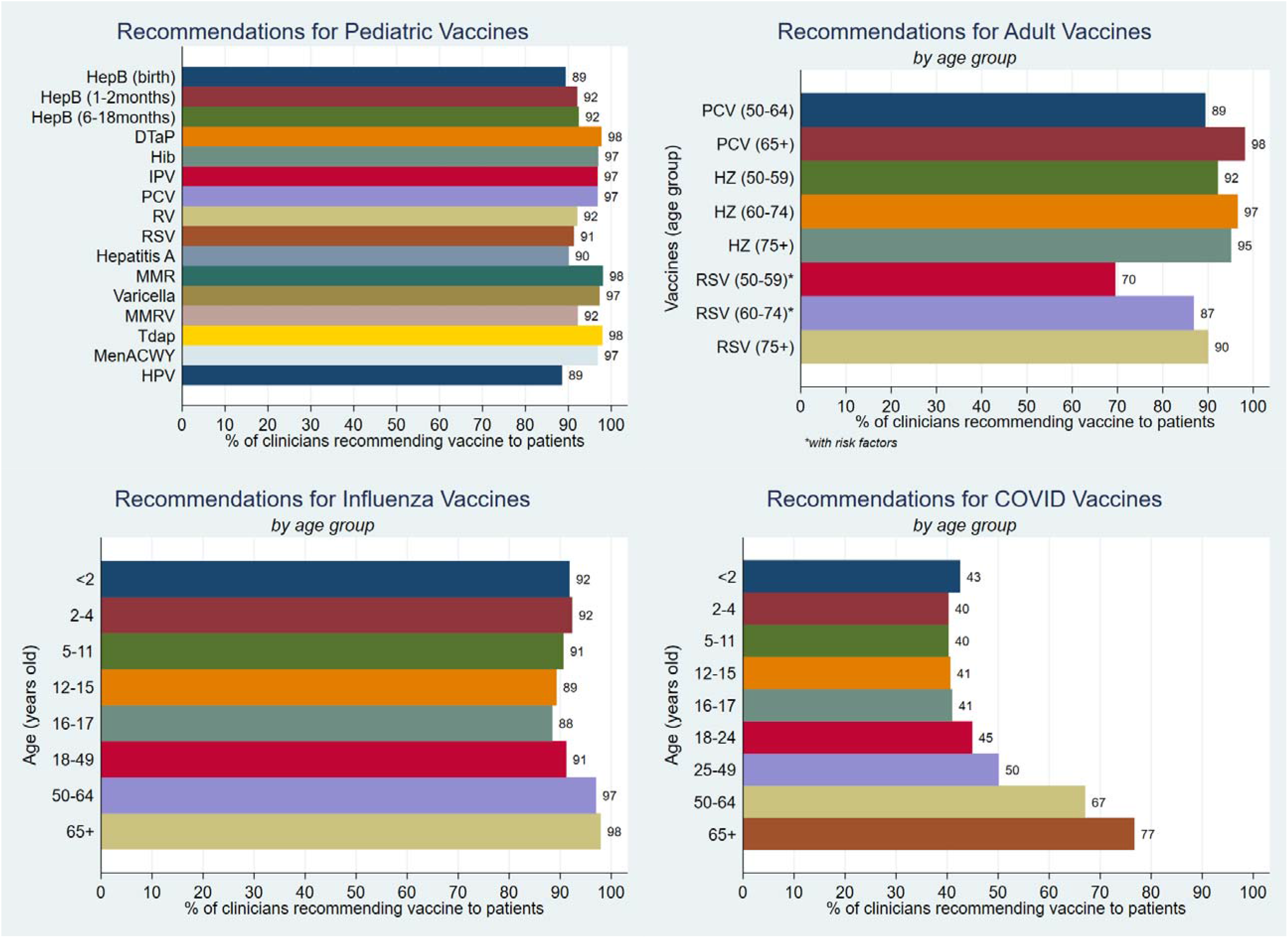
Proportion of Clinicians Recommending Specific Vaccines for Patients. Note: Respondents were asked whether they provided care for children, young adults, and/or older adults, and were only presented survey items pertaining to vaccines relevant for their patient populations; percentages presented in figures refer to clinicians recommending or strongly recommending the vaccine. Vaccine acronyms: DTaP = Diphtheria, Tetanus, and Acellular Pertussis (pediatric); HepB = Hepatitis B; Hib = Haemophilus influenzae type b; HPV = Human Papillomavirus; IPV = Inactivated Poliovirus Vaccine; MenACWY = Meningococcal Conjugate Vaccine; MMR = Measles, Mumps, and Rubella; MMRV = Measles, Mumps, Rubella, and Varicella; PCV = Pneumococcal Conjugate Vaccine; RV = Rotavirus; RSV = Respiratory syncytial virus; Tdap = Tetanus, Diphtheria, and Acellular Pertussis (adult booster); HZ = Herpes Zoster

Clinician recommendations for human papillomavirus (HPV) vaccine varied widely by patient age. While 87% of clinicians recommended vaccination at 11-12 years of age and 93-94% of clinicians recommended vaccination at 13-17 years of age, only 60% recommended vaccination at 9-10 years of age, despite AAP’s recommendation to initiate HPV vaccination as early as age 9 (Supplemental Figure 2).^23^

Seasonal influenza vaccination was recommended by most clinicians, whether for children (91-92%), adolescents (88-89%), young adults (91%), or older adults (97-98%) (Figure 2). Most clinicians (89-98%) also recommended routine vaccination of older adults with herpes zoster, PCV, and RSV vaccines. In contrast, less than half of clinicians recommended COVID-19 vaccination of infants (43%), children (40%), adolescents (41%), or young adults (45-50%), though nearly two-thirds (67%) recommended vaccination of adults 50-64 years of age, and more than three-quarters (77%) recommended vaccination of adults at least 65 years of age. Strong adult vaccine recommendations were most common for patients at least 65 years of age to receive influenza vaccine (95%) and PCV (93%) (Supplemental Table 5).

### Vaccine Discussions with Patients

More than half (54%) of clinicians reported that at least one-quarter of their patients expressed concerns about vaccines (Supplemental Table 6). While most (88%) clinicians felt well prepared for vaccine conversations with patients, nearly one-quarter (23%) felt they did not have everything they needed to share vaccine information with their patients. Nearly one-quarter (22%) of clinicians found it difficult to stay up to date on vaccine recommendations, and nearly half (46%) found it difficult to stay up to date on vaccine concerns. Nearly three-quarters (71%) reported that their patients sometimes asked vaccine questions to which they did not know the answer. More than one-quarter (26%) of clinicians reported never billing insurance for vaccine counseling.

More than three-quarters of clinicians reported increases over the past year (i.e., since June 2025) in the frequency (80%) and intensity (76%) of patient vaccine concerns, implying a substantial increase in both the number of concerned patients and the level of concern among patients (Supplemental Figure 3). Many clinicians also reported spending more time talking with patients about vaccines (61%) and looking up vaccine information (45%).

### Patient Vaccine Refusal

Clinician-estimated patient refusal rates were typically lower for pediatric vaccines than for adult vaccines (Supplemental Table 7) (Supplemental Figures 4 and 5). Clinicians reported a high patient vaccine refusal rate (i.e., more than 10%) most frequently for adult (91%) and pediatric (89%) COVID-19, adult (83%) and pediatric (82%) influenza, herpes zoster (73%), HPV (71%), adult (75%) and pediatric (50%) RSV, adult PCV (65%), and the hepatitis B birth dose (42%) (Figure 3).

**Figure 3.**
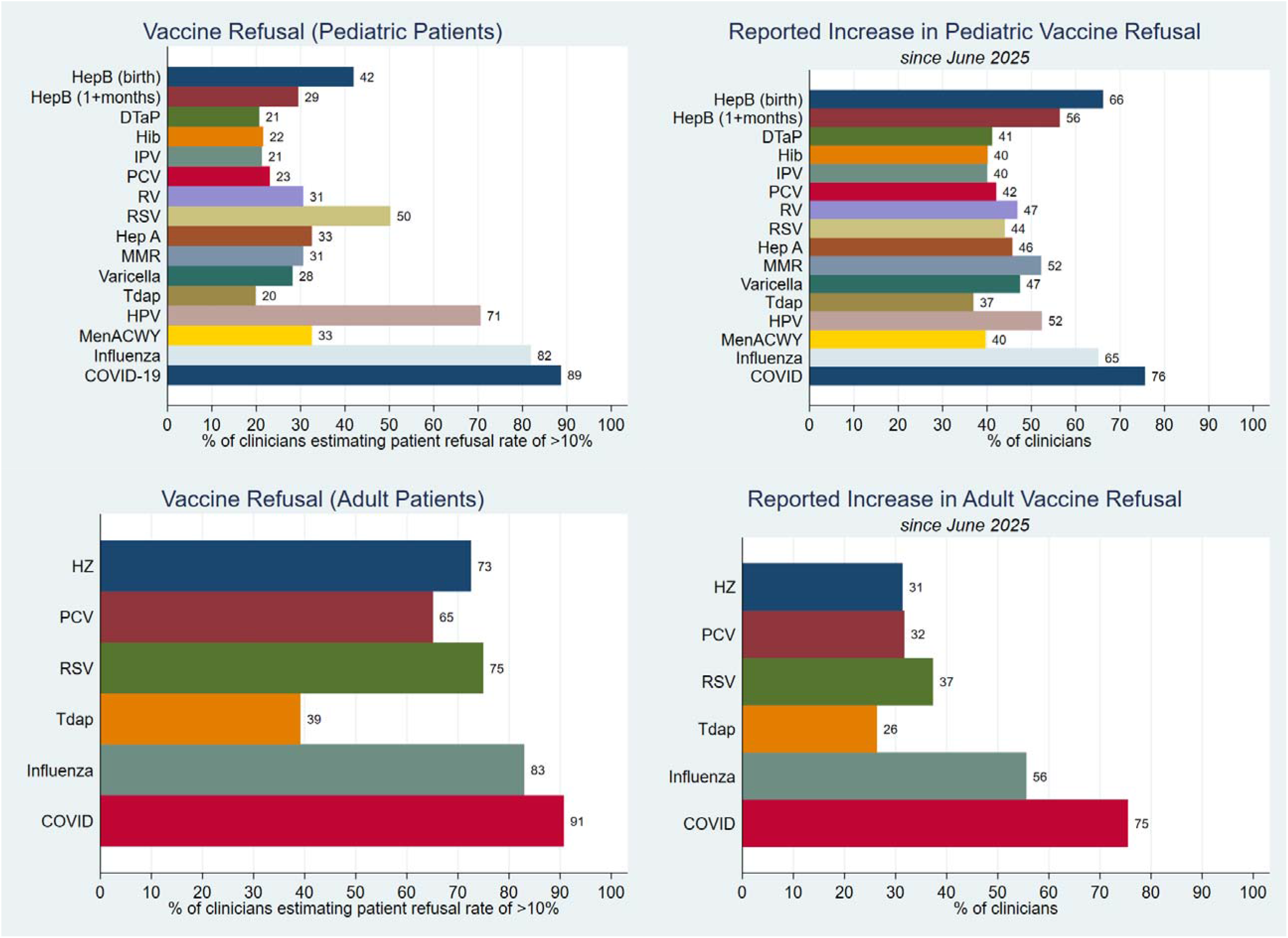
Reported Rates of and Increases in Patient Vaccine Refusal from June 2025 to July 2026. Vaccine acronyms: DTaP = Diphtheria, Tetanus, and Acellular Pertussis (pediatric); HepB = Hepatitis B; Hib = Haemophilus influenzae type b; HPV = Human Papillomavirus; IPV = Inactivated Poliovirus Vaccine; MenACWY = Meningococcal Conjugate Vaccine; MMR = Measles, Mumps, and Rubella; MMRV = Measles, Mumps, Rubella, and Varicella; PCV = Pneumococcal Conjugate Vaccine; RV = Rotavirus; RSV = Respiratory syncytial virus; Tdap = Tetanus, Diphtheria, and Acellular Pertussis (adult booster); HZ = Herpes Zoster

Increases in patient refusal rates since June 2025 were reported by three-quarters of clinicians for both pediatric (76%) and adult (75%) COVID-19 vaccination, by nearly two-thirds of clinicians for hepatitis B at birth (66%) and pediatric influenza vaccination (65%), and by more than half of clinicians for MMR (52%), HPV (52%), hepatitis B at one month (56%), and adult influenza (56%) (Figure 3). Decreases in patient refusal were reported most frequently for RSV (10%) and HPV (8%) among pediatric vaccines, and for PCV (11%), RSV (11%), and shingles (9%) among adult vaccines (Supplemental Table 8).

### Differences by Trust in CDC Vaccine Recommendations

Less than one third (30%) of pediatricians trusted current CDC vaccine recommendations, compared to about half of PAs (49%) and family physicians (51%), and nearly two-thirds of NPs (65%) and nurses (66%) (p<0.01) (Table 1). Clinicians who voted for Donald Trump in the 2024 presidential election were far more likely to trust current CDC vaccine recommendations than clinicians who voted for Kamala Harris (61% vs 33%, p<0.01).

Clinicians who trusted current CDC vaccine recommendations were less likely to recommend hepatitis B vaccines, whether at birth (87% vs 91%, p=0.01), at 1-2 months (90% vs 94%, p=0.02), or at 6-18 months (91% vs 94%, p=0.04) (Supplemental Table 9). Other pediatric vaccines recommended less frequently by clinicians who trusted CDC vaccine recommendations included hepatitis A (86% vs 93%, p<0.01), rotavirus (89% vs 95%, p<0.01), and RSV (89% vs 93%, p=0.04). Clinicians who trusted current CDC vaccine recommendations were also less likely to recommend HPV vaccine for their 11-12-year-old patients (84% vs 90%, p=0.02) and COVID-19 vaccine for their 6-23-month-old patients (38% vs 46%, p<0.01). Additionally, clinicians who trusted current CDC vaccine recommendations were less likely to report increases since June 2025 in the frequency (74% vs 86%) and intensity (71% vs 81%) of patient vaccine concerns (p<0.01), the amount of time spent talking with patients about vaccines (53% vs 68%, p<0.01), and patient refusal across all pediatric and several adult vaccines (Supplemental Table 10).

### Changes Since 2023

Compared to January 2023, fewer clinicians in July 2026 reported frequently obtaining vaccine information from news media, websites, journal publications, blogs, message boards, text message alerts, and podcasts, and fewer clinicians reported trusting vaccine information from news media, other clinicians, health departments, and federal agencies (adjusted p≤0.01) (Table 2). The sources of vaccine information with the largest declines in clinician trust over this period by far were CDC (47% vs 79%, adjusted p<0.01) and FDA (43% vs 75%, adjusted p<0.01). Likewise, far fewer clinicians reported trust in CDC generally (46% vs 74%, adjusted p<0.01). While clinician recommendations for most vaccines have remained stable since 2023, more clinicians in 2026 reported recommending shingles vaccine to their patients, and fewer clinicians reported recommending COVID-19 vaccines across all age groups (adjusted p<0.01) (Table 2).

**Table 2.**
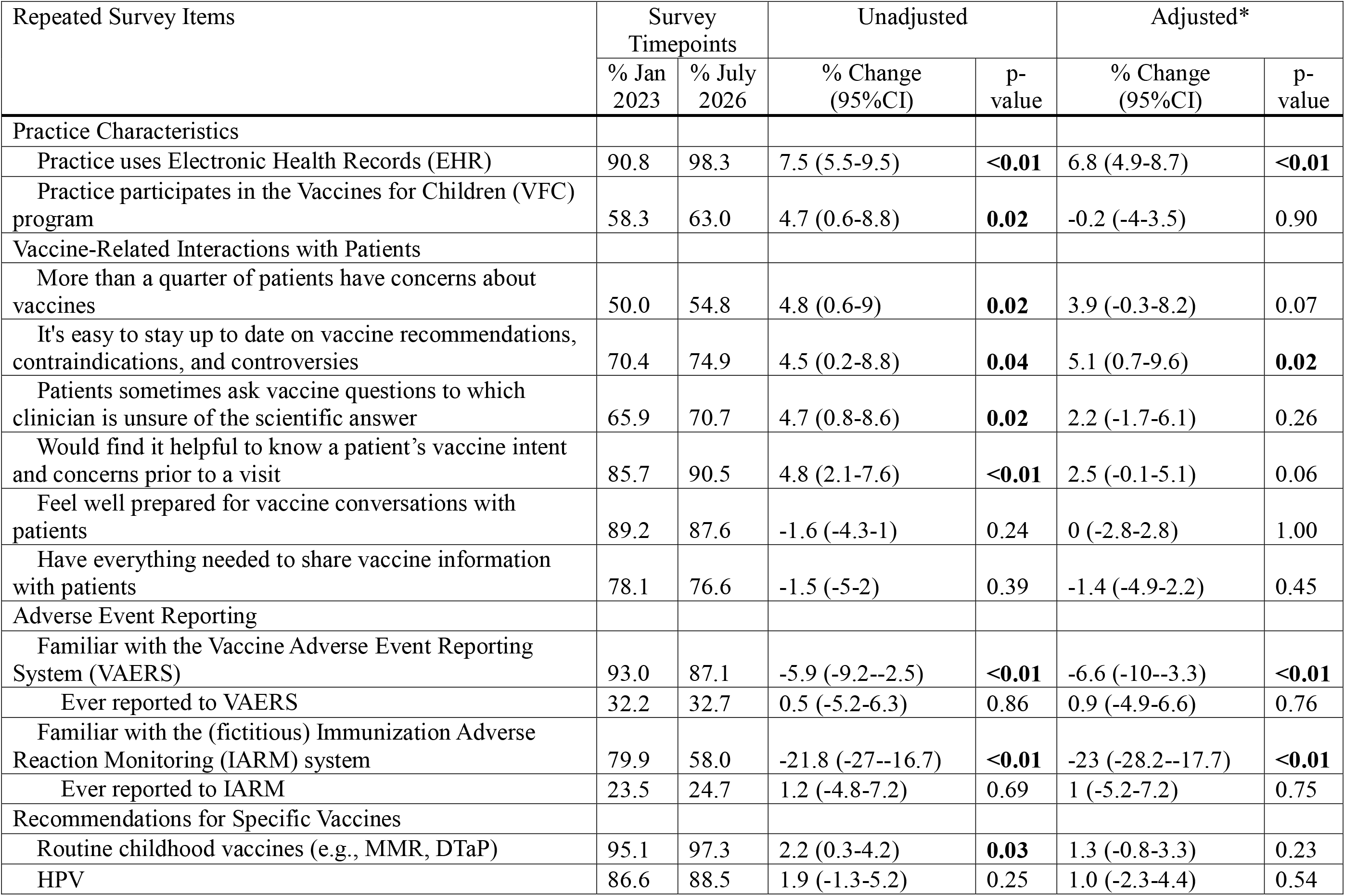
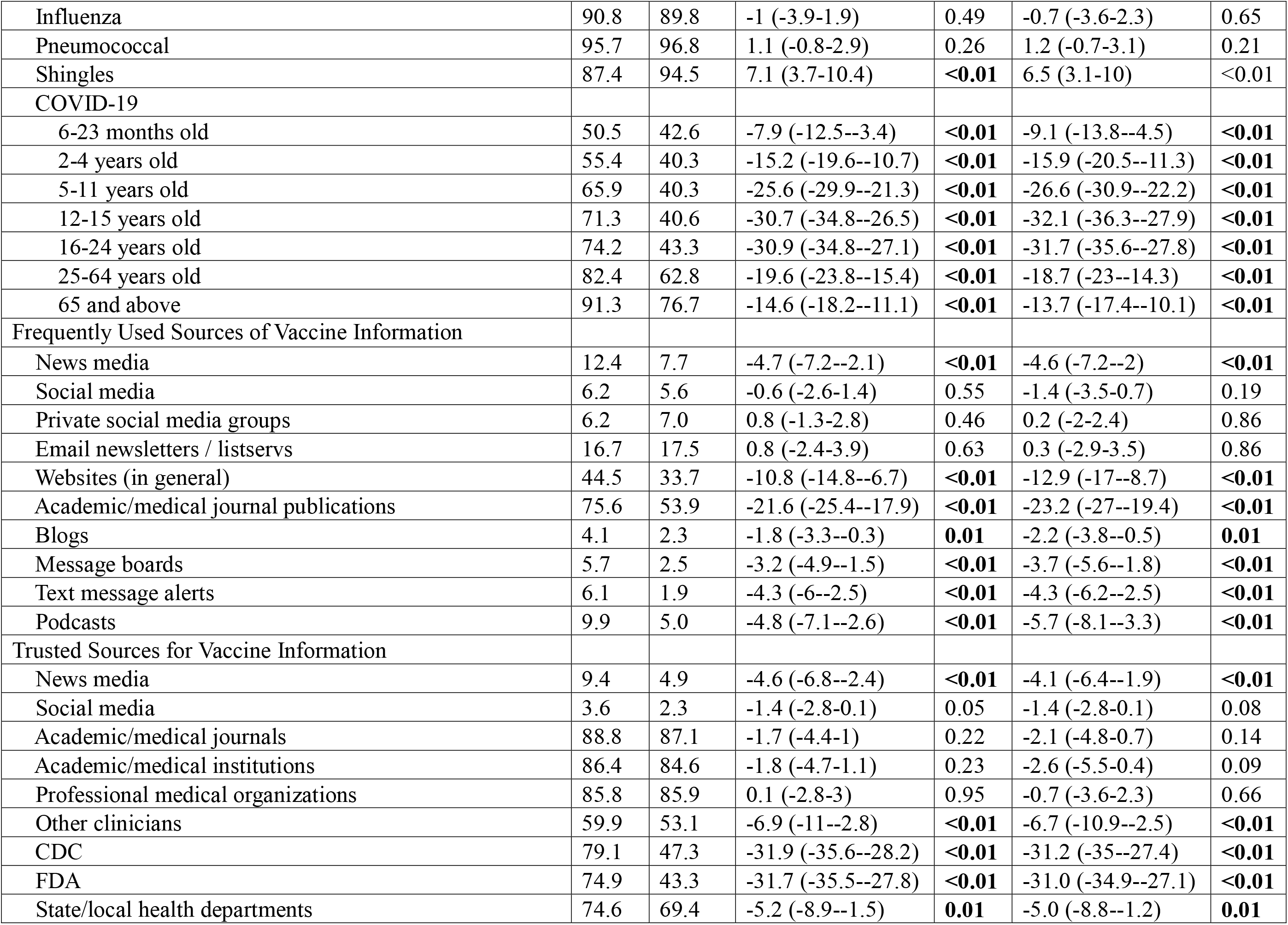
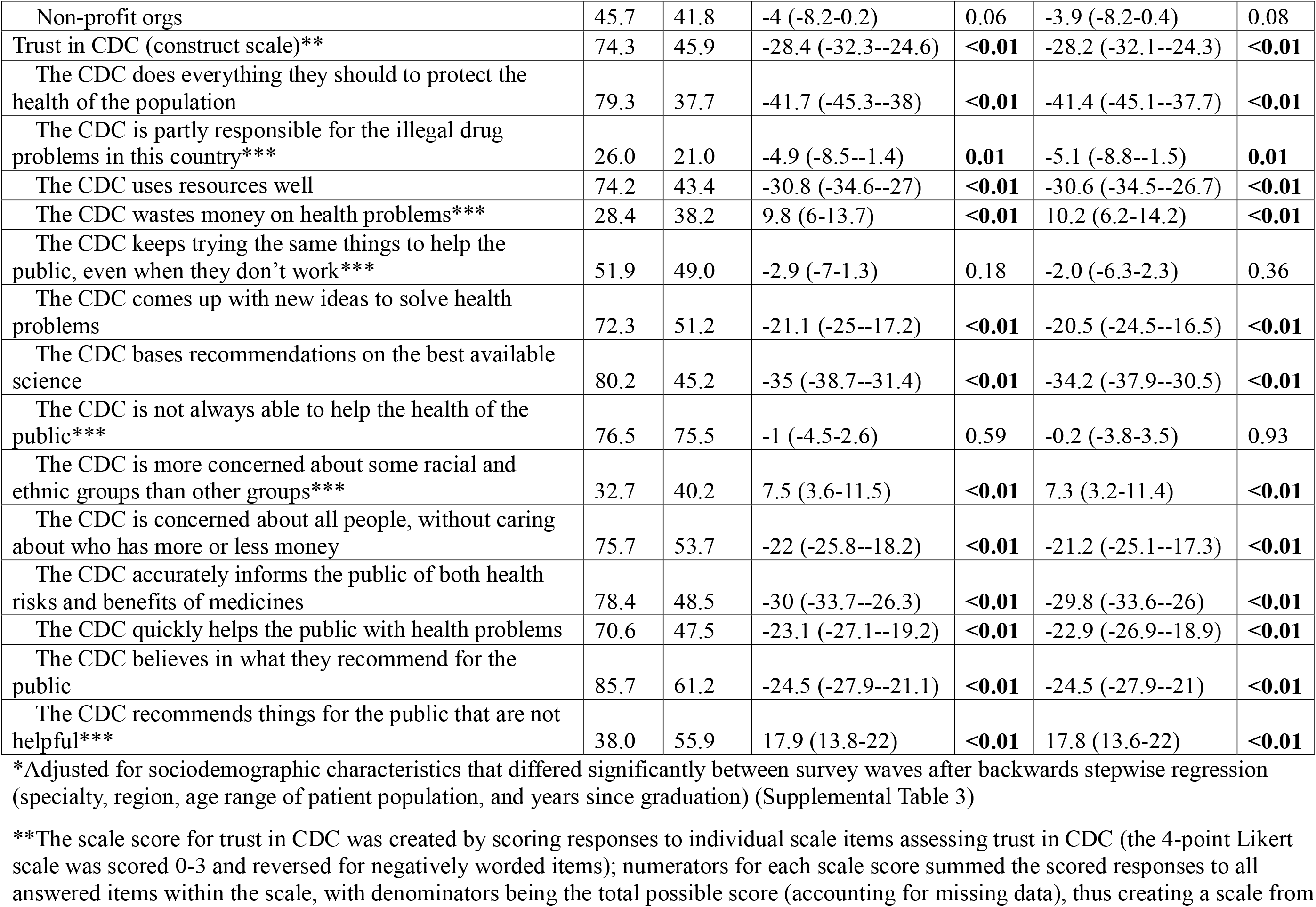
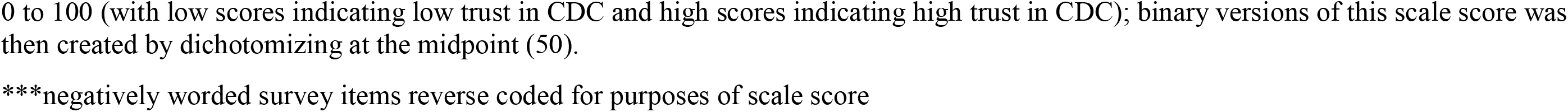
Changes in Clinician Responses between January 2023 and July 2026.

| Repeated Survey Items | Survey Timepoints |  | Unadjusted |  | Adjusted* |  |
|---|---|---|---|---|---|---|
|  | % Jan 2023 | % July 2026 | % Change (95%CI) | p-value | % Change (95%CI) | p-value |
| Practice Characteristics |  |  |  |  |  |  |
| Practice uses Electronic Health Records (EHR) | 90.8 | 98.3 | 7.5 (5.5-9.5) | **<0.01** | 6.8 (4.9-8.7) | **<0.01** |
| Practice participates in the Vaccines for Children (VFC) program | 58.3 | 63.0 | 4.7 (0.6-8.8) | **0.02** | -0.2 (-4-3.5) | 0.90 |
| Vaccine-Related Interactions with Patients |  |  |  |  |  |  |
| More than a quarter of patients have concerns about vaccines | 50.0 | 54.8 | 4.8 (0.6-9) | **0.02** | 3.9 (-0.3-8.2) | 0.07 |
| It's easy to stay up to date on vaccine recommendations, contraindications, and controversies | 70.4 | 74.9 | 4.5 (0.2-8.8) | **0.04** | 5.1 (0.7-9.6) | **0.02** |
| Patients sometimes ask vaccine questions to which clinician is unsure of the scientific answer | 65.9 | 70.7 | 4.7 (0.8-8.6) | **0.02** | 2.2 (-1.7-6.1) | 0.26 |
| Would find it helpful to know a patient’s vaccine intent and concerns prior to a visit | 85.7 | 90.5 | 4.8 (2.1-7.6) | **<0.01** | 2.5 (-0.1-5.1) | 0.06 |
| Feel well prepared for vaccine conversations with patients | 89.2 | 87.6 | -1.6 (-4.3-1) | 0.24 | 0 (-2.8-2.8) | 1.00 |
| Have everything needed to share vaccine information with patients | 78.1 | 76.6 | -1.5 (-5-2) | 0.39 | -1.4 (-4.9-2.2) | 0.45 |
| Adverse Event Reporting |  |  |  |  |  |  |
| Familiar with the Vaccine Adverse Event Reporting System (VAERS) | 93.0 | 87.1 | -5.9 (-9.2--2.5) | **<0.01** | -6.6 (-10--3.3) | **<0.01** |
| Ever reported to VAERS | 32.2 | 32.7 | 0.5 (-5.2-6.3) | 0.86 | 0.9 (-4.9-6.6) | 0.76 |
| Familiar with the (fictitious) Immunization Adverse Reaction Monitoring (IARM) system | 79.9 | 58.0 | -21.8 (-27--16.7) | **<0.01** | -23 (-28.2--17.7) | **<0.01** |
| Ever reported to IARM | 23.5 | 24.7 | 1.2 (-4.8-7.2) | 0.69 | 1 (-5.2-7.2) | 0.75 |
| Recommendations for Specific Vaccines |  |  |  |  |  |  |
| Routine childhood vaccines (e.g., MMR, DTaP) | 95.1 | 97.3 | 2.2 (0.3-4.2) | **0.03** | 1.3 (-0.8-3.3) | 0.23 |
| HPV | 86.6 | 88.5 | 1.9 (-1.3-5.2) | 0.25 | 1.0 (-2.3-4.4) | 0.54 |
| Influenza | 90.8 | 89.8 | -1 (-3.9-1.9) | 0.49 | -0.7 (-3.6-2.3) | 0.65 |
| Pneumococcal | 95.7 | 96.8 | 1.1 (-0.8-2.9) | 0.26 | 1.2 (-0.7-3.1) | 0.21 |
| Shingles | 87.4 | 94.5 | 7.1 (3.7-10.4) | **<0.01** | 6.5 (3.1-10) | <0.01 |
| COVID-19 |  |  |  |  |  |  |
| 6-23 months old | 50.5 | 42.6 | -7.9 (-12.5--3.4) | **<0.01** | -9.1 (-13.8--4.5) | **<0.01** |
| 2-4 years old | 55.4 | 40.3 | -15.2 (-19.6--10.7) | **<0.01** | -15.9 (-20.5--11.3) | **<0.01** |
| 5-11 years old | 65.9 | 40.3 | -25.6 (-29.9--21.3) | **<0.01** | -26.6 (-30.9--22.2) | **<0.01** |
| 12-15 years old | 71.3 | 40.6 | -30.7 (-34.8--26.5) | **<0.01** | -32.1 (-36.3--27.9) | **<0.01** |
| 16-24 years old | 74.2 | 43.3 | -30.9 (-34.8--27.1) | **<0.01** | -31.7 (-35.6--27.8) | **<0.01** |
| 25-64 years old | 82.4 | 62.8 | -19.6 (-23.8--15.4) | **<0.01** | -18.7 (-23--14.3) | **<0.01** |
| 65 and above | 91.3 | 76.7 | -14.6 (-18.2--11.1) | **<0.01** | -13.7 (-17.4--10.1) | **<0.01** |
| Frequently Used Sources of Vaccine Information |  |  |  |  |  |  |
| News media | 12.4 | 7.7 | -4.7 (-7.2--2.1) | **<0.01** | -4.6 (-7.2--2) | **<0.01** |
| Social media | 6.2 | 5.6 | -0.6 (-2.6-1.4) | 0.55 | -1.4 (-3.5-0.7) | 0.19 |
| Private social media groups | 6.2 | 7.0 | 0.8 (-1.3-2.8) | 0.46 | 0.2 (-2-2.4) | 0.86 |
| Email newsletters / listservs | 16.7 | 17.5 | 0.8 (-2.4-3.9) | 0.63 | 0.3 (-2.9-3.5) | 0.86 |
| Websites (in general) | 44.5 | 33.7 | -10.8 (-14.8--6.7) | **<0.01** | -12.9 (-17--8.7) | **<0.01** |
| Academic/medical journal publications | 75.6 | 53.9 | -21.6 (-25.4--17.9) | **<0.01** | -23.2 (-27--19.4) | **<0.01** |
| Blogs | 4.1 | 2.3 | -1.8 (-3.3--0.3) | **0.01** | -2.2 (-3.8--0.5) | **0.01** |
| Message boards | 5.7 | 2.5 | -3.2 (-4.9--1.5) | **<0.01** | -3.7 (-5.6--1.8) | **<0.01** |
| Text message alerts | 6.1 | 1.9 | -4.3 (-6--2.5) | **<0.01** | -4.3 (-6.2--2.5) | **<0.01** |
| Podcasts | 9.9 | 5.0 | -4.8 (-7.1--2.6) | **<0.01** | -5.7 (-8.1--3.3) | **<0.01** |
| Trusted Sources for Vaccine Information |  |  |  |  |  |  |
| News media | 9.4 | 4.9 | -4.6 (-6.8--2.4) | **<0.01** | -4.1 (-6.4--1.9) | **<0.01** |
| Social media | 3.6 | 2.3 | -1.4 (-2.8-0.1) | 0.05 | -1.4 (-2.8-0.1) | 0.08 |
| Academic/medical journals | 88.8 | 87.1 | -1.7 (-4.4-1) | 0.22 | -2.1 (-4.8-0.7) | 0.14 |
| Academic/medical institutions | 86.4 | 84.6 | -1.8 (-4.7-1.1) | 0.23 | -2.6 (-5.5-0.4) | 0.09 |
| Professional medical organizations | 85.8 | 85.9 | 0.1 (-2.8-3) | 0.95 | -0.7 (-3.6-2.3) | 0.66 |
| Other clinicians | 59.9 | 53.1 | -6.9 (-11--2.8) | **<0.01** | -6.7 (-10.9--2.5) | **<0.01** |
| CDC | 79.1 | 47.3 | -31.9 (-35.6--28.2) | **<0.01** | -31.2 (-35--27.4) | **<0.01** |
| FDA | 74.9 | 43.3 | -31.7 (-35.5--27.8) | **<0.01** | -31.0 (-34.9--27.1) | **<0.01** |
| State/local health departments | 74.6 | 69.4 | -5.2 (-8.9--1.5) | **0.01** | -5.0 (-8.8--1.2) | **0.01** |
| Non-profit orgs | 45.7 | 41.8 | -4 (-8.2-0.2) | 0.06 | -3.9 (-8.2-0.4) | 0.08 |
| Trust in CDC (construct scale)** | 74.3 | 45.9 | -28.4 (-32.3--24.6) | **<0.01** | -28.2 (-32.1--24.3) | **<0.01** |
| The CDC does everything they should to protect the health of the population | 79.3 | 37.7 | -41.7 (-45.3--38) | **<0.01** | -41.4 (-45.1--37.7) | **<0.01** |
| The CDC is partly responsible for the illegal drug problems in this country*** | 26.0 | 21.0 | -4.9 (-8.5--1.4) | **0.01** | -5.1 (-8.8--1.5) | **0.01** |
| The CDC uses resources well | 74.2 | 43.4 | -30.8 (-34.6--27) | **<0.01** | -30.6 (-34.5--26.7) | **<0.01** |
| The CDC wastes money on health problems*** | 28.4 | 38.2 | 9.8 (6-13.7) | **<0.01** | 10.2 (6.2-14.2) | **<0.01** |
| The CDC keeps trying the same things to help the public, even when they don’t work*** | 51.9 | 49.0 | -2.9 (-7-1.3) | 0.18 | -2.0 (-6.3-2.3) | 0.36 |
| The CDC comes up with new ideas to solve health problems | 72.3 | 51.2 | -21.1 (-25--17.2) | **<0.01** | -20.5 (-24.5--16.5) | **<0.01** |
| The CDC bases recommendations on the best available science | 80.2 | 45.2 | -35 (-38.7--31.4) | **<0.01** | -34.2 (-37.9--30.5) | **<0.01** |
| The CDC is not always able to help the health of the public*** | 76.5 | 75.5 | -1 (-4.5-2.6) | 0.59 | -0.2 (-3.8-3.5) | 0.93 |
| The CDC is more concerned about some racial and ethnic groups than other groups*** | 32.7 | 40.2 | 7.5 (3.6-11.5) | **<0.01** | 7.3 (3.2-11.4) | **<0.01** |
| The CDC is concerned about all people, without caring about who has more or less money | 75.7 | 53.7 | -22 (-25.8--18.2) | **<0.01** | -21.2 (-25.1--17.3) | **<0.01** |
| The CDC accurately informs the public of both health risks and benefits of medicines | 78.4 | 48.5 | -30 (-33.7--26.3) | **<0.01** | -29.8 (-33.6--26) | **<0.01** |
| The CDC quickly helps the public with health problems | 70.6 | 47.5 | -23.1 (-27.1--19.2) | **<0.01** | -22.9 (-26.9--18.9) | **<0.01** |
| The CDC believes in what they recommend for the public | 85.7 | 61.2 | -24.5 (-27.9--21.1) | **<0.01** | -24.5 (-27.9--21) | **<0.01** |
| The CDC recommends things for the public that are not helpful*** | 38.0 | 55.9 | 17.9 (13.8-22) | **<0.01** | 17.8 (13.6-22) | **<0.01** |

\*Adjusted for sociodemographic characteristics that differed significantly between survey waves after backwards stepwise regression (specialty, region, age range of patient population, and years since graduation) (Supplemental Table 3)
\*\*The scale score for trust in CDC was created by scoring responses to individual scale items assessing trust in CDC (the 4-point Likert scale was scored 0-3 and reversed for negatively worded items); numerators for each scale score summed the scored responses to all answered items within the scale, with denominators being the total possible score (accounting for missing data), thus creating a scale from 0 to 100 (with low scores indicating low trust in CDC and high scores indicating high trust in CDC); binary versions of this scale score was then created by dichotomizing at the midpoint (50).
\*\*\*negatively worded survey items reverse coded for purposes of scale score

## Discussion

This large, high-quality panel survey demonstrates continued strong support of vaccination from most clinicians despite recent fluctuations in federal vaccine recommendations. However, many clinicians reported turning to nongovernmental sources of vaccine information and recommendations as their trust in federal agencies diminished. Reports of increased patient vaccine hesitancy and time spent counseling patients about vaccines reflect both the burden on clinicians and the potential impact of vaccine policy changes on patients and their families.

Despite concerns that volatility in federal vaccine recommendations may affect clinician vaccine confidence, the only vaccine for which clinician recommendations clearly decreased over time was COVID-19. Although AAP and AAFP have maintained a universal recommendation for COVID-19 vaccination of children 6-23 months of age,^24,25^ fewer than half of pediatric clinicians adhere to this recommendation. Similarly, while AAFP continues to recommend all adults receive a COVID-19 vaccine annually,^25^ fewer than half of clinicians recommend that their young adult patients be vaccinated. Reduced support of COVID-19 vaccination may partly reflect AAP’s recent discontinuation of their universal recommendation for children 2-18 years of age,^24^ rather than federal policy changes. It may also reflect trends predating 2025 such as the politicization of COVID-19 vaccines or general pandemic fatigue.^26–28^

While clinician recommendations for other vaccines have remained stable, many clinicians reported increases since June 2025 in vaccine refusal (especially for COVID-19, influenza, and the hepatitis B birth dose), suggesting that changes in federal recommendations and the corresponding media discourse may have undermined public confidence in vaccines. Serial, representative public surveys are needed to monitor fluctuations in vaccine confidence.

Increased patient concerns appear to have increased the time clinicians spend discussing vaccines with patients and looking up vaccine information. Most clinicians have limited time, and reimbursement for vaccine administration does not adequately compensate clinicians for counseling patients about vaccines.^29–31^ While some billing codes are available for vaccine counseling, clinicians rarely use these codes, potentially because their reimbursement from insurance companies is inconsistent.^32–34^ Improved reimbursement would allow clinicians to spend more time discussing vaccines with patients without financial repercussions. Additionally, infodemiology could be leveraged to identify emerging concerns and prepare clinicians accordingly.^35^

Historically, clinicians have had high trust in CDC, facilitating federal efforts to address infectious disease through vaccination.^19^ Recent events have clearly undermined clinician trust in CDC and other federal agencies. To reverse this decline, federal officials should consider returning to processes for vaccine recommendations that prioritize expertise, professional society representation, and transparent policymaking. Professional associations have become clinicians’ primary source of vaccine information and recommendations, giving them the opportunity to play a greater role in supporting clinician vaccine knowledge and confidence.

The main limitation of this survey is its lack of a true probability sample, meaning it remains prone to selection and self-selection biases and cannot be assumed to represent all U.S. clinicians. However, this sample was derived from a high-quality panel and used quotas to ensure broad participation across targeted specialties and regions. Other limitations arise from the survey’s length and reliance on clinician self-reporting. The multitude of survey items enables diverse analyses but increases the risk of false positive findings due to multiple comparisons. Self-reported data is prone to social desirability and recall biases, exemplified by the many clinicians who reported trust in vaccine information from NVIC (a nonprofit organization criticized for misrepresenting scientific evidence on vaccines)^36^ and awareness of the IARM system (a fictitious version of VAERS that we use to test for these biases). Still, clinician reports of patient vaccine attitudes and behavior can serve as an early warning in lieu of true vaccine coverage data, which are much harder to obtain in a timely manner. Coverage estimates from the CDC’s National Immunization Survey often take years to reach the scientific and medical communities,^37^ and annual rates of exemptions to school immunization requirements largely reflect parents’ vaccine decisions made years earlier (when their children were the recommended age for routine vaccination),^38^ so neither can be relied on to capture current fluctuations in vaccine confidence and uptake. Additionally, while coverage data directly reflect parental vaccine decision-making, there is added value in measuring the upstream input of clinician vaccine recommendations and experiences. However, the three-year gap between this survey and the most recent comparison survey makes it difficult to determine a precise turning point in clinician responses or connect fluctuations with specific events. Future surveys should be conducted with more regularity to enable such determinations.

This large, high-quality panel survey provides insight into the current vaccine-related attitudes and experiences of U.S. clinicians as well as the point-of-care impact of the recent upheaval in federal vaccine policy. Professional associations can support immunization efforts by providing clinicians with reliable vaccine information and recommendations.

## Data Availability

The deidentified survey dataset will be available from the corresponding author upon reasonable request following the publication of planned secondary analyses.

## Acknowledgments

We would like to acknowledge with gratitude the individuals who shared their invaluable feedback during the design and revision of the survey instrument, including Dr. Walter Orenstein (Professor Emeritus, Emory University), Dr. Michelle Fiscus (Chief Medical Officer, Association of Immunization Managers), Dr. Douglas Opel (Professor, Department of Pediatrics, University of Washington School of Medicine), Dr. Kawsar Talaat (Associate Professor, Department of International Health, Johns Hopkins Bloomberg School of Public Health), and Ms. Tina Proveaux (Communications Specialist, Department of International Health, Johns Hopkins Bloomberg School of Public Health).

## Author Contributions

Dr. Dudley drafted, revised, and finalized the survey instrument; performed all data management and analysis; and drafted, revised, and finalized this manuscript. Dr. Sharfstein, Dr. Moss, Dr. Weeks, Ms. Melchinger, Ms. Goryn, and Dr. Salmon reviewed and provided feedback on the survey instrument. All coauthors contributed to interpretation of the data and reviewed and provided feedback and final approval on this manuscript.

Dr. Dudley had full access to all the data in the study and takes responsibility for the integrity of the data and the accuracy of the data analysis.

## Supplemental Materials

**Supplemental Table 1.** Recruitment Data for July 2026 Clinician Survey.

|  | **Total** | **Family Physicians** | **Pediatricians** | **Nurses** | **NPs** | **PAs** |
|---|---|---|---|---|---|---|
| **Invites sent** | **13990** | **3674** | **2447** | **1343** | **4749** | **1777** |
| **Total responses** | **3032** | **883** | **787** | **351** | **746** | **265** |
| **Response rate** | **22%** | **24%** | **32%** | **26%** | **16%** | **15%** |
| **Excluded from sample** | **1522** | **383** | **279** | **185** | **580** | **98** |
| Over quota (region and/or clinician type) | 593 | 177 | 129 | 7 | 280 | 0 |
| Incomplete (screener) | 598 | 158 | 129 | 62 | 176 | 73 |
| Incomplete (main survey) | 45 | 7 | 8 | 21 | 7 | 2 |
| Terminated as ineligible | 274 | 38 | 10 | 92 | 113 | 21 |
| *S1b - Other Specialty (Nurses/NPs/PAs)* | 171 | 0 | 0 | 78 | 79 | 14 |
| *S2 - No Direct Patient Care* | 19 | 7 | 7 | 2 | 3 | 0 |
| *S3 - No Vaccines Administered* | 84 | 31 | 3 | 12 | 31 | 7 |
| Quality Removals (e.g., straight liners)* | 12 | 3 | 3 | 3 | 4 | 2 |
| **Included in final sample** | **1510** | **500** | **508** | **166** | **166** | **167** |

\* straight liners refers to clinicians who chose the same response for an entire survey section (e.g., choosing "strongly agree" for all 14 items in the trust in CDC scale, despite some items describing CDC positively and some items describing CDC negatively)

**Supplemental Table 2.** Comparison of Clinicians in Panel Responding vs Not Responding to Survey Invitation.

|  | Total<br>N (%)* | Responders<br>N (%)* | Non-responders<br>N (%)* | p-value |
|---|---|---|---|---|
| All | N=13,990 | 3,032 (22%) | 10,958 (78%) |  |
| **Clinician Characteristics** |  |  |  |  |
| Clinician Type |  |  |  | **<0.01** |
| Pediatrician | 2447 (17%) | 954 (31%) | 1493 (14%) |  |
| Family Physician | 3674 (26%) | 1044 (34%) | 2630 (24%) |  |
| Nurse Practitioner (NP) | 4749 (34%) | 324 (11%) | 4425 (40%) |  |
| Physician Assistant (PA) | 1777 (13%) | 277 (9%) | 1500 (14%) |  |
| Nurse | 1343 (10%) | 433 (14%) | 910 (8%) |  |
| Region |  |  |  | **<0.01** |
| Midwest | 2632 (19%) | 556 (18%) | 2157 (20%) |  |
| Northeast | 2713 (19%) | 539 (18%) | 2093 (19%) |  |
| South | 5530 (40%) | 879 (29%) | 4651 (42%) |  |
| West | 2376 (17%) | 600 (20%) | 1776 (16%) |  |
| Unknown | 739 (5%) | 459 (15%) | 280 (3%) |  |
| Gender |  |  |  | **<0.01** |
| Male | 3981 (28%) | 1542 (51%) | 6775 (62%) |  |
| Female | 8317 (59%) | 1464 (48%) | 2517 (23%) |  |
| Non-binary | 54 (0%) | 4 (0%) | 50 (0%) |  |
| Unknown | 1638 (12%) | 22 (1%) | 1616 (15%) |  |

\* Column percentages

**Supplemental Table 3.** Differences in Sample Characteristics between January 2023 and July 2026 Surveys.

|  | % Jan 2023 | % July 2026 | % Change (95%CI) | p-value |
|---|---|---|---|---|
| Clinician Characteristics |  |  |  |  |
| Clinician Type |  |  |  | 1.00 |
| Pediatrician | 33.7 | 33.7 | 0 (-3.9-3.9) |  |
| Family Physician | 33.2 | 33.5 | 0.3 (-3.6-4.2) |  |
| Nurse Practitioner (NP) | 11.0 | 11.0 | 0 (-2.6-2.6) |  |
| Physician Assistant (PA) | 11.1 | 10.6 | -0.5 (-3.1-2) |  |
| Nurse | 11.0 | 11.2 | 0.2 (-2.4-2.8) |  |
| Specialty |  |  |  | **<0.01** |
| Pediatrics | 41.3 | 37.2 | -4 (-8.1-0) |  |
| Family Practice | 53.7 | 50.6 | -3.1 (-7.3-1) |  |
| Internal Medicine | 5.0 | 12.1 | 7.2 (4.7-9.6) |  |
| Highest completed clinical degree |  |  |  | 0.75 |
| Associate | 1.7 | 2.5 | 0.8 (-0.5-2) |  |
| Bachelor's | 7.2 | 7.0 | -0.1 (-2.3-2) |  |
| Master's | 21.2 | 20.2 | -1 (-4.3-2.4) |  |
| Doctoral | 69.7 | 70.0 | 0.3 (-3.5-4.1) |  |
| Other | 0.2 | 0.2 | 0 (-0.4-0.4) |  |
| Years since graduation |  |  |  | **<0.01** |
| <10 | 29.9 | 14.4 | -15.5 (-18.8--12.2) |  |
| 10-19 | 29.6 | 26.6 | -3 (-6.8-0.7) |  |
| 20-29 | 21.9 | 30.8 | 8.9 (5.2-12.6) |  |
| 30-39 | 13.0 | 20.0 | 7 (3.8-10.1) |  |
| 40+ | 5.6 | 8.3 | 2.7 (0.6-4.9) |  |
| Region |  |  |  | **<0.01** |
| Midwest | 23.2 | 27.0 | 3.9 (0.2-7.5) |  |
| Northeast | 22.4 | 24.2 | 1.8 (-1.7-5.3) |  |
| South | 31.4 | 31.4 | 0.1 (-3.8-3.9) |  |
| West | 23.1 | 17.4 | -5.7 (-9--2.4) |  |
| Race/Ethnicity |  |  |  | 0.72 |
| Asian | 14.8 | 13.7 | -1.1 (-4.1-1.9) |  |
| Black | 4.0 | 4.8 | 0.8 (-1-2.6) |  |
| Hispanic/Latino | 4.2 | 3.4 | -0.8 (-2.4-0.9) |  |
| White | 73.9 | 74.8 | 0.9 (-2.9-4.6) |  |
| Other/Multiple | 3.2 | 3.4 | 0.2 (-1.3-1.8) |  |
| Practice Characteristics |  |  |  |  |
| Practice Setting |  |  |  | **<0.01** |
| Private Practice | 50.5 | 54.8 | 4.3 (0.2-8.5) |  |
| Practice Network or Health Maintenance Organization (HMO) | 15.1 | 11.8 | -3.3 (-6.1--0.5) |  |
| Hospital or Medical Center | 21.2 | 24.9 | 3.7 (0.2-7.2) |  |
| Community Health Center or Federally Qualified Health Center (FQHC) | 11.1 | 6.7 | -4.4 (-6.7--2.1) |  |
| Other | 2.1 | 1.8 | -0.3 (-1.5-0.8) |  |
| Urbanicity of Practice Location |  |  |  | 0.30 |
| Rural | 16.0 | 13.7 | -2.3 (-5.2-0.7) |  |
| Suburban | 48.0 | 50.2 | 2.1 (-2-6.3) |  |
| Urban | 36.0 | 36.1 | 0.1 (-3.9-4.1) |  |
| Age Range of Patient Population |  |  |  | **<0.01** |
| Children only | 22.9 | 21.5 | -1.4 (-4.9-2) |  |
| Adults only | 11.0 | 15.8 | 4.8 (1.9-7.6) |  |
| Both Children & Adults | 66.1 | 62.8 | -3.3 (-7.3-0.7) |  |

**Supplemental Table 4.** Patient Populations of Participating Clinicians, Stratified by Trust in CDC Vaccine Recommendations.

|  | Total<br>N (%)* | Trust in CDC Vaccine Recommendations |  | p-value |
|---|---|---|---|---|
|  |  | Low Trust<br>N (%)** | High Trust<br>N (%)** |  |
| All | N=1,507 | 801 (53.2%) | 704 (46.8%) |  |
| Age Range of Patient Population |  |  |  | **<0.01** |
| Children only | 345 (22.9%) | 226 (65.5%) | 119 (34.5%) |  |
| Young Adults (18-49) only | 10 (0.7%) | 7 (70.0%) | 3 (30.0%) |  |
| Older Adults (50+) only | 17 (1.1%) | 6 (35.3%) | 11 (64.7%) |  |
| Children & Young Adults only | 278 (18.4%) | 184 (66.2%) | 94 (33.8%) |  |
| Young Adults & Older Adults only | 139 (9.2%) | 49 (35.3%) | 90 (64.7%) |  |
| All | 718 (47.6%) | 329 (45.9%) | 387 (54.1%) |  |
| Race/Ethnicity of Patient Population |  |  |  |  |
| Hispanic or Latino |  |  |  | 0.12 |
| <25% | 1,071 (71.1%) | 581 (54.3%) | 488 (45.7%) |  |
| 25-50% | 360 (23.9%) | 175 (48.6%) | 185 (51.4%) |  |
| 51-75% | 54 (3.6%) | 34 (63.0%) | 20 (37.0%) |  |
| >75% | 22 (1.5%) | 11 (50.0%) | 11 (50.0%) |  |
| Black or African American |  |  |  | 0.06 |
| <25% | 1,069 (70.9%) | 592 (55.4%) | 476 (44.6%) |  |
| 25-50% | 395 (26.2%) | 189 (48.0%) | 205 (52.0%) |  |
| 51-75% | 34 (2.3%) | 15 (44.1%) | 19 (55.9%) |  |
| >75% | 9 (0.6%) | 5 (55.6%) | 4 (44.4%) |  |
| Asian |  |  |  | 0.14 |
| <25% | 1,413 (93.8%) | 742 (52.6%) | 669 (47.4%) |  |
| 25-50% | 84 (5.6%) | 51 (60.7%) | 33 (39.3%) |  |
| 51-75% | 8 (0.5%) | 6 (75.0%) | 2 (25.0%) |  |
| >75% | 2 (0.1%) | 2 (100.0%) | 0 ( 0.0%) |  |
| White |  |  |  | 0.44 |
| <25% | 216 (14.3%) | 116 (53.7%) | 100 (46.3%) |  |
| 25-50% | 567 (37.6%) | 287 (50.6%) | 280 (49.4%) |  |
| 51-75% | 463 (30.7%) | 252 (54.7%) | 209 (45.3%) |  |
| >75% | 261 (17.3%) | 146 (55.9%) | 115 (44.1%) |  |
| Health Insurance Status of Patient Population |  |  |  |  |
| Private Insurance |  |  |  | **<0.01** |
| <25% | 239 (15.9%) | 117 (49.2%) | 121 (50.8%) |  |
| 25-50% | 611 (40.5%) | 301 (49.3%) | 310 (50.7%) |  |
| 51-75% | 420 (27.9%) | 235 (56.0%) | 185 (44.0%) |  |
| >75% | 237 (15.7%) | 148 (62.7%) | 88 (37.3%) |  |
| Medicaid or CHIP |  |  |  | 0.15 |
| <25% | 763 (50.6%) | 396 (52.0%) | 365 (48.0%) |  |
| 25-50% | 529 (35.1%) | 280 (52.9%) | 249 (47.1%) |  |
| 51-75% | 149 (9.9%) | 81 (54.4%) | 68 (45.6%) |  |
| >75% | 66 (4.4%) | 44 (66.7%) | 22 (33.3%) |  |
| Medicare |  |  |  | **<0.01** |
| <25% | 954 (63.3%) | 555 (58.3%) | 397 (41.7%) |  |
| 25-50% | 505 (33.5%) | 231 (45.7%) | 274 (54.3%) |  |
| 51-75% | 34 (2.3%) | 11 (32.4%) | 23 (67.6%) |  |
| >75% | 14 (0.9%) | 4 (28.6%) | 10 (71.4%) |  |
| Uninsured |  |  |  | **0.02** |
| <25% | 1,451 (96.3%) | 781 (53.9%) | 669 (46.1%) |  |
| 25-50% | 51 (3.4%) | 17 (33.3%) | 34 (66.7%) |  |
| 51-75% | 3 (0.2%) | 2 (100.0%) | 0 ( 0.0%) |  |
| >75% | 2 (0.1%) | 1 (50.0%) | 1 (50.0%) |  |

\* Column percentages out of total sample (N=1,507)
\*\*Row percentages among subsample with data on trust in CDC vaccine recommendations (N=1,505); N=2 survey respondents reported being unfamiliar with CDC vaccine recommendations and were thus counted as missing for this variable

**Supplemental Table 5.** Strength of Clinician Vaccine Recommendations for Patients.

| Specific Vaccines | Strongly Recommend Against N (%)* | Recommend Against N (%)* | Neutral N (%)* | Recommend N (%)* | Strongly Recommend N (%)* |
|---|---|---|---|---|---|
| Hepatitis B |  |  |  |  |  |
| At birth | 42 ( 3.5%) | 17 ( 1.4%) | 69 ( 5.7%) | 114 ( 9.5%) | 960 (79.9%) |
| At 1-2 months old | 40 ( 3.2%) | 17 ( 1.4%) | 42 ( 3.4%) | 104 ( 8.3%) | 1,045 (83.7%) |
| At 6-18 months old | 37 ( 2.9%) | 10 ( 0.8%) | 49 ( 3.9%) | 105 ( 8.3%) | 1,069 (84.2%) |
| Routine childhood vaccines (e.g., MMR, DTaP)<sup>2</sup> | 12 ( 1.8%) | 1 ( 0.2%) | 5 ( 0.8%) | 17 ( 2.6%) | 627 (94.7%) |
| DTaP<sup>1</sup> | 7 ( 1.1%) | 1 ( 0.2%) | 7 ( 1.1%) | 18 ( 2.7%) | 629 (95.0%) |
| Hib<sup>1</sup> | 7 ( 1.1%) | 1 ( 0.2%) | 12 ( 1.8%) | 20 ( 3.0%) | 621 (93.9%) |
| IPV<sup>1</sup> | 6 ( 0.9%) | 3 ( 0.5%) | 12 ( 1.8%) | 28 ( 4.2%) | 611 (92.6%) |
| Pneumococcal<sup>1</sup> | 8 ( 1.2%) | 1 ( 0.2%) | 12 ( 1.8%) | 28 ( 4.2%) | 614 (92.6%) |
| RV<sup>1</sup> | 8 ( 1.2%) | 14 ( 2.1%) | 30 ( 4.6%) | 73 (11.1%) | 531 (80.9%) |
| RSV (monoclonal antibody)<sup>1</sup> | 7 ( 1.1%) | 8 ( 1.2%) | 42 ( 6.4%) | 92 (14.0%) | 507 (77.3%) |
| Hepatitis A<sup>1</sup> | 9 ( 1.4%) | 15 ( 2.3%) | 42 ( 6.3%) | 90 (13.5%) | 509 (76.5%) |
| MMR<sup>1</sup> | 7 ( 1.0%) | 1 ( 0.1%) | 5 ( 0.7%) | 17 ( 2.5%) | 637 (95.5%) |
| Varicella<sup>1</sup> | 7 ( 1.1%) | 1 ( 0.2%) | 10 ( 1.5%) | 29 ( 4.4%) | 619 (92.9%) |
| MMRV<sup>1</sup> | 17 ( 2.6%) | 11 ( 1.7%) | 24 ( 3.6%) | 33 ( 5.0%) | 581 (87.2%) |
| Routine adolescent vaccines |  |  |  |  |  |
| Tdap<sup>1</sup> | 8 ( 1.2%) | 1 ( 0.1%) | 5 ( 0.7%) | 19 ( 2.8%) | 638 (95.1%) |
| MenACWY<sup>1</sup> | 8 ( 1.2%) | 2 ( 0.3%) | 11 ( 1.6%) | 53 ( 7.9%) | 597 (89.0%) |
| HPV (all ages)<sup>2</sup> | 10 ( 1.4%) | 13 ( 1.8%) | 61 ( 8.3%) | 121 (16.5%) | 527 (72.0%) |
| 9-10 years old<sup>1</sup> | 24 ( 3.6%) | 47 ( 7.1%) | 194 (29.2%) | 129 (19.4%) | 271 (40.8%) |
| 11-12 years old<sup>1</sup> | 10 ( 1.5%) | 18 ( 2.7%) | 58 ( 8.7%) | 111 (16.6%) | 471 (70.5%) |
| 13-15 years old<sup>1</sup> | 11 ( 1.6%) | 5 ( 0.7%) | 25 ( 3.7%) | 90 (13.4%) | 540 (80.5%) |
| 16-17 years old<sup>1</sup> | 10 ( 1.5%) | 4 ( 0.6%) | 30 ( 4.5%) | 81 (12.0%) | 548 (81.4%) |
| 18-26 years old<sup>1</sup> | 8 ( 1.4%) | 10 ( 1.8%) | 41 ( 7.2%) | 109 (19.1%) | 402 (70.5%) |
| 27-45 years old<sup>1</sup> | 12 ( 2.4%) | 37 ( 7.5%) | 146 (29.4%) | 112 (22.6%) | 189 (38.1%) |
| Respiratory vaccines |  |  |  |  |  |
| Influenza (all ages)<sup>2</sup> | 7 ( 0.9%) | 9 ( 1.2%) | 61 ( 8.1%) | 197 (26.1%) | 480 (63.7%) |
| 6-23 months old<sup>1</sup> | 5 ( 0.8%) | 8 ( 1.2%) | 40 ( 6.2%) | 98 (15.1%) | 498 (76.7%) |
| 2-4 years old<sup>1</sup> | 3 ( 0.5%) | 14 ( 2.1%) | 33 ( 5.1%) | 109 (16.7%) | 494 (75.7%) |
| 5-11 years old<sup>1</sup> | 5 ( 0.8%) | 9 ( 1.4%) | 48 ( 7.2%) | 134 (20.2%) | 467 (70.4%) |
| 12-15 years old<sup>1</sup> | 5 ( 0.7%) | 14 ( 2.1%) | 53 ( 7.9%) | 145 (21.6%) | 454 (67.7%) |
| 16-17 years old<sup>1</sup> | 5 ( 0.7%) | 14 ( 2.1%) | 59 ( 8.7%) | 142 (21.0%) | 455 (67.4%) |
| 18-49 years old<sup>1</sup> | 4 ( 0.7%) | 5 ( 0.9%) | 41 ( 7.2%) | 125 (22.0%) | 392 (69.1%) |
| 50-64 years old<sup>1</sup> | 4 ( 0.9%) | 0 ( 0.0%) | 9 ( 2.1%) | 56 (12.8%) | 367 (84.2%) |
| 65 and above<sup>1</sup> | 3 ( 0.7%) | 1 ( 0.2%) | 5 ( 1.2%) | 15 ( 3.5%) | 409 (94.5%) |
| Pneumococcal (all ages)<sup>2</sup> | 8 ( 1.1%) | 2 ( 0.3%) | 14 ( 1.9%) | 76 (10.2%) | 647 (86.6%) |
| 50-64 years old<sup>1</sup> | 4 ( 0.9%) | 3 ( 0.7%) | 39 ( 9.0%) | 120 (27.6%) | 268 (61.8%) |
| 65 and above<sup>1</sup> | 3 ( 0.7%) | 0 ( 0.0%) | 5 ( 1.2%) | 24 ( 5.5%) | 402 (92.6%) |
| RSV |  |  |  |  |  |
| 50-59 years old (with risk factor) | 13 ( 1.5%) | 53 ( 6.1%) | 199 (22.9%) | 237 (27.3%) | 367 (42.2%) |
| 60-74 years old (with risk factor) | 6 ( 0.7%) | 19 ( 2.2%) | 89 (10.2%) | 206 (23.7%) | 549 (63.2%) |
| 75 and above | 6 ( 0.7%) | 17 ( 2.0%) | 63 ( 7.3%) | 161 (18.7%) | 616 (71.4%) |
| COVID-19 |  |  |  |  |  |
| 6-23 months old | 131 (10.2%) | 218 (17.0%) | 388 (30.2%) | 232 (18.1%) | 314 (24.5%) |
| 2-4 years old | 120 ( 9.3%) | 230 (17.8%) | 420 (32.6%) | 226 (17.5%) | 293 (22.7%) |
| 5-11 years old | 116 ( 8.8%) | 227 (17.3%) | 440 (33.6%) | 229 (17.5%) | 299 (22.8%) |
| 12-15 years old | 115 ( 8.7%) | 227 (17.1%) | 447 (33.6%) | 234 (17.6%) | 306 (23.0%) |
| 16-17 years old | 111 ( 8.3%) | 227 (17.0%) | 452 (33.8%) | 239 (17.8%) | 310 (23.2%) |
| 18-24 years old | 91 ( 8.0%) | 180 (15.8%) | 357 (31.3%) | 228 (20.0%) | 284 (24.9%) |
| 25-49 years old | 71 ( 7.1%) | 137 (13.6%) | 294 (29.2%) | 217 (21.6%) | 287 (28.5%) |
| 50-64 years old | 40 ( 4.6%) | 68 ( 7.8%) | 179 (20.6%) | 227 (26.1%) | 357 (41.0%) |
| 65 and above | 34 ( 3.9%) | 48 ( 5.5%) | 121 (13.9%) | 170 (19.5%) | 498 (57.2%) |
| Other routine adult vaccines |  |  |  |  |  |
| Shingles (all ages)<sup>2</sup> | 2 ( 0.5%) | 1 ( 0.2%) | 21 ( 4.8%) | 101 (23.2%) | 311 (71.3%) |
| 50-59 years old<sup>1</sup> | 4 ( 0.9%) | 1 ( 0.2%) | 29 ( 6.7%) | 82 (18.9%) | 319 (73.3%) |
| 60-74 years old<sup>1</sup> | 3 ( 0.7%) | 0 ( 0.0%) | 12 ( 2.8%) | 53 (12.2%) | 368 (84.4%) |
| 75 and above<sup>1</sup> | 3 ( 0.7%) | 1 ( 0.2%) | 17 ( 3.9%) | 72 (16.7%) | 338 (78.4%) |

\*Respondents were asked whether they provided care for children, young adults, and/or older adults, and were only presented survey items pertaining to vaccines relevant for their patient populations; half of respondents were randomized to receive items asking about their recommendations for specific vaccines and age groups<sup>1</sup>, while the other half received items asking about their vaccine recommendations more generally<sup>2</sup> (to facilitate direct comparison with a previous survey).<sup>19</sup>

**Supplemental Table 6.** Clinician-Patient Vaccine-Related Interactions, Stratified by Trust in CDC Vaccine Recommendations.

|  | Total<br>N (%)* | Trust in CDC Vaccine Recommendations |  | p-value |
|---|---|---|---|---|
|  |  | Low Trust<br>N (%)** | Low Trust<br>N (%)** |  |
| Patient Vaccine Hesitancy |  |  |  |  |
| Pediatric patients with parent(s) concerned about vaccines |  |  |  | 0.94 |
| 0% | 12 (0.9%) | 6 (50.0%) | 6 (50.0%) |  |
| 1-24% | 603 (45.0%) | 338 (56.1%) | 264 (43.9%) |  |
| 25-50% | 539 (40.2%) | 296 (54.9%) | 243 (45.1%) |  |
| 51-75% | 128 (9.5%) | 67 (52.3%) | 61 (47.7%) |  |
| 76-100% | 59 (4.4%) | 32 (55.2%) | 26 (44.8%) |  |
| Adult patients concerned about vaccines |  |  |  | 0.27 |
| 0% | 25 (2.2%) | 17 (68.0%) | 8 (32.0%) |  |
| 1-24% | 512 (44.1%) | 247 (48.2%) | 265 (51.8%) |  |
| 25-50% | 470 (40.4%) | 233 (49.8%) | 235 (50.2%) |  |
| 51-75% | 117 (10.1%) | 62 (53.0%) | 55 (47.0%) |  |
| 76-100% | 38 (3.3%) | 16 (42.1%) | 22 (57.9%) |  |
| Hours spent per week discussing vaccines with patients |  |  |  | **<0.01** |
| <1 | 280 (18.7%) | 114 (41.0%) | 164 (59.0%) |  |
| 1 | 351 (23.4%) | 187 (53.3%) | 164 (46.7%) |  |
| 2-3 | 483 (32.2%) | 288 (59.6%) | 195 (40.4%) |  |
| 4-5 | 210 (14.0%) | 125 (59.5%) | 85 (40.5%) |  |
| 6-9 | 71 (4.7%) | 38 (53.5%) | 33 (46.5%) |  |
| 10+ | 105 (7.0%) | 47 (44.8%) | 58 (55.2%) |  |
| Minutes spent per clinical encounter discussing vaccines with patients |  |  |  | **<0.01** |
| 0-1 | 73 (5.0%) | 39 (53.4%) | 34 (46.6%) |  |
| 2-3 | 442 (30.1%) | 260 (58.8%) | 182 (41.2%) |  |
| 4-5 | 605 (41.2%) | 327 (54.2%) | 276 (45.8%) |  |
| 6-10 | 256 (17.4%) | 124 (48.4%) | 132 (51.6%) |  |
| 11-19 | 49 (3.3%) | 16 (32.7%) | 33 (67.3%) |  |
| 20+ | 45 (3.1%) | 22 (48.9%) | 23 (51.1%) |  |
| Minutes spent per week looking up vaccine information |  |  |  | **<0.01** |
| 0 | 100 (6.6%) | 67 (67.0%) | 33 (33.0%) |  |
| 1-3 | 125 (8.3%) | 66 (52.8%) | 59 (47.2%) |  |
| 4-5 | 240 (15.9%) | 144 (60.5%) | 94 (39.5%) |  |
| 6-10 | 255 (16.9%) | 140 (54.9%) | 115 (45.1%) |  |
| 11-19 | 125 (8.3%) | 61 (48.8%) | 64 (51.2%) |  |
| 20-29 | 108 (7.2%) | 56 (51.9%) | 52 (48.1%) |  |
| 30-59 | 207 (13.7%) | 96 (46.4%) | 111 (53.6%) |  |
| 60-99 | 216 (14.3%) | 121 (56.0%) | 95 (44.0%) |  |
| 100+ | 131 (8.7%) | 50 (38.2%) | 81 (61.8%) |  |
| It's easy to stay up to date on vaccine recommendations and contraindications | 585 (77.6%) | 300 (74.6%) | 283 (80.9%) | **0.04** |
| It's easy to stay up to date on vaccine concerns and controversies | 479 (63.5%) | 246 (61.2%) | 231 (66.0%) | 0.17 |
| Patients sometimes ask vaccine questions to which clinician is unsure of the scientific answer | 1,065 (70.7%) | 553 (69.0%) | 510 (72.4%) | 0.15 |
| Would find it helpful to know patient’s vaccine intent and concerns prior to visit | 1,364 (90.5%) | 719 (89.8%) | 644 (91.5%) | 0.26 |
| Feel well prepared for vaccine conversations with patients | 1,320 (87.6%) | 698 (87.1%) | 620 (88.1%) | 0.59 |
| Have everything needed to share vaccine information with patients | 1,154 (76.6%) | 615 (76.8%) | 537 (76.3%) | 0.82 |
| Concerned about liability for administering vaccines to patients | 168 (11.1%) | 67 ( 8.4%) | 101 (14.3%) | **<0.01** |
| Method of Obtaining Informed Consent for Vaccination |  |  |  |  |
| Written/signed consent | 788 (52.3%) | 389 (48.6%) | 398 (56.5%) | **<0.01** |
| Verbal/oral consent | 938 (62.2%) | 526 (65.7%) | 411 (58.4%) | **<0.01** |
| Digital/electronic consent | 348 (23.1%) | 159 (19.9%) | 189 (26.8%) | **<0.01** |
| Implied consent | 126 (8.4%) | 70 ( 8.7%) | 56 ( 8.0%) | 0.58 |
| Billing for Vaccination |  |  |  |  |
| % of visits with vaccine given that vaccine administration billed for |  |  |  | **<0.01** |
| None | 39 (2.6%) | 24 (63.2%) | 14 (36.8%) |  |
| <25% | 96 (6.4%) | 33 (34.4%) | 63 (65.6%) |  |
| 25-50% | 110 (7.3%) | 47 (43.1%) | 62 (56.9%) |  |
| 51-75% | 89 (5.9%) | 32 (36.0%) | 57 (64.0%) |  |
| >75% | 874 (58.0%) | 508 (58.1%) | 366 (41.9%) |  |
| Unsure | 299 (19.8%) | 157 (52.5%) | 142 (47.5%) |  |
| % of visits with vaccine given that vaccine counseling billed for |  |  |  | 0.05 |
| None | 385 (25.5%) | 199 (51.8%) | 185 (48.2%) |  |
| <25% | 245 (16.3%) | 121 (49.4%) | 124 (50.6%) |  |
| 25-50% | 108 (7.2%) | 53 (49.1%) | 55 (50.9%) |  |
| 51-75% | 83 (5.5%) | 37 (44.6%) | 46 (55.4%) |  |
| >75% | 308 (20.4%) | 185 (60.1%) | 123 (39.9%) |  |
| Unsure | 378 (25.1%) | 206 (54.6%) | 171 (45.4%) |  |
| % of visits with vaccine discussed but not given that vaccine counseling billed for |  |  |  | 0.09 |
| None | 564 (37.4%) | 304 (54.0%) | 259 (46.0%) |  |
| <25% | 335 (22.2%) | 182 (54.3%) | 153 (45.7%) |  |
| 25-50% | 117 (7.8%) | 53 (45.3%) | 64 (54.7%) |  |
| 51-75% | 63 (4.2%) | 30 (47.6%) | 33 (52.4%) |  |
| >75% | 103 (6.8%) | 66 (64.1%) | 37 (35.9%) |  |
| Unsure | 325 (21.6%) | 166 (51.2%) | 158 (48.8%) |  |
| Use of Vaccine Information Statements (VIS) |  |  |  |  |
| % of visits with vaccine given that VIS provided to patient |  |  |  | **<0.01** |
| None | 51 (3.4%) | 18 (35.3%) | 33 (64.7%) |  |
| <25% | 117 (7.8%) | 55 (47.0%) | 62 (53.0%) |  |
| 25-50% | 100 (6.6%) | 41 (41.4%) | 58 (58.6%) |  |
| 51-75% | 93 (6.2%) | 45 (48.4%) | 48 (51.6%) |  |
| >75% | 1,032 (68.5%) | 594 (57.6%) | 438 (42.4%) |  |
| Unsure | 114 (7.6%) | 48 (42.5%) | 65 (57.5%) |  |
| Adverse Event Reporting |  |  |  |  |
| Familiar with the Vaccine Adverse Event Reporting System (VAERS) | 657 (87.1%) | 366 (92.2%) | 291 (81.5%) | **<0.01** |
| Ever reported to VAERS | 215 (32.7%) | 128 (35.0%) | 87 (29.9%) | 0.17 |
| Familiar with the (fictitious) Immunization Adverse Reaction Monitoring (IARM) system | 437 (58.0%) | 238 (58.9%) | 197 (56.8%) | 0.55 |
| Ever reported to (fictitious) IARM system | 108 (24.7%) | 65 (27.3%) | 42 (21.3%) | 0.15 |
| Perceived Utility of Resources to Improve Vaccine-Related Interactions with Patients |  |  |  |  |
| For Clinicians |  |  |  |  |
| An in-person training on evidence-based strategies for vaccine discussions with patients (with CME credit) | 642 (42.6%) | 272 (34.0%) | 369 (52.4%) | **<0.01** |
| An online training module on evidence-based strategies for vaccine discussions with patients (with CME credit) | 853 (56.6%) | 403 (50.3%) | 449 (63.8%) | **<0.01** |
| A guide summarizing scientific literature on vaccine safety concerns | 998 (66.2%) | 489 (61.0%) | 508 (72.2%) | **<0.01** |
| Guidance on billing and reimbursement for vaccine counseling | 791 (52.5%) | 395 (49.3%) | 395 (56.1%) | **<0.01** |
| Legal advice on vaccine-related liability | 604 (40.1%) | 282 (35.2%) | 322 (45.7%) | **<0.01** |
| For Patients |  |  |  |  |
| An educational website with personalized videos about vaccines | 932 (61.8%) | 447 (55.8%) | 483 (68.6%) | **<0.01** |
| A chatbot that answers vaccine questions | 620 (41.1%) | 292 (36.5%) | 327 (46.4%) | **<0.01** |
| Printable handouts on vaccines | 868 (57.6%) | 396 (49.4%) | 470 (66.8%) | **<0.01** |

\* Column percentages out of total sample (N=1,507)
\*\*Row percentages among subsample with data on trust in CDC vaccine recommendations (N=1,505); N=2 survey respondents reported being unfamiliar with CDC vaccine recommendations and were thus counted as missing for this variable

**Supplemental Table 7.** Estimated Patient Vaccine Refusal Rate from June 2025 to July 2026.

|  | Estimated Patient Refusal Rate |  |  |  |  |  |  |  |
|---|---|---|---|---|---|---|---|---|
|  | 0% | 1-5% | 6-10% | 11-20% | 21-30% | 31-40% | 41-50% | 51-100% |
| Pediatric Vaccines* |  |  |  |  |  |  |  |  |
| Hepatitis B |  |  |  |  |  |  |  |  |
| At birth | 184 (13.7%) | 324 (24.2%) | 271 (20.2%) | 232 (17.3%) | 138 (10.3%) | 46 ( 3.4%) | 77 ( 5.7%) | 69 ( 5.1%) |
| At 1+ months old | 198 (14.8%) | 456 (34.0%) | 292 (21.8%) | 194 (14.5%) | 90 ( 6.7%) | 29 ( 2.2%) | 40 ( 3.0%) | 42 ( 3.1%) |
| DTaP | 271 (20.2%) | 523 (39.0%) | 270 (20.1%) | 139 (10.4%) | 61 ( 4.5%) | 14 ( 1.0%) | 25 ( 1.9%) | 38 ( 2.8%) |
| Hib | 279 (20.8%) | 520 (38.8%) | 253 (18.9%) | 145 (10.8%) | 69 ( 5.1%) | 9 ( 0.7%) | 29 ( 2.2%) | 37 ( 2.8%) |
| IPV | 290 (21.6%) | 527 (39.3%) | 239 (17.8%) | 149 (11.1%) | 60 ( 4.5%) | 10 ( 0.7%) | 27 ( 2.0%) | 39 ( 2.9%) |
| Pneumococcal | 267 (19.9%) | 500 (37.3%) | 265 (19.8%) | 151 (11.3%) | 68 ( 5.1%) | 18 ( 1.3%) | 28 ( 2.1%) | 44 ( 3.3%) |
| RV | 216 (16.1%) | 454 (33.9%) | 261 (19.5%) | 206 (15.4%) | 80 ( 6.0%) | 31 ( 2.3%) | 34 ( 2.5%) | 59 ( 4.4%) |
| RSV | 175 (13.0%) | 276 (20.6%) | 217 (16.2%) | 209 (15.6%) | 160 (11.9%) | 76 ( 5.7%) | 121 ( 9.0%) | 107 ( 8.0%) |
| Hepatitis A | 197 (14.7%) | 437 (32.6%) | 271 (20.2%) | 192 (14.3%) | 97 ( 7.2%) | 30 ( 2.2%) | 53 ( 4.0%) | 64 ( 4.8%) |
| MMR | 203 (15.1%) | 454 (33.9%) | 274 (20.4%) | 212 (15.8%) | 91 ( 6.8%) | 35 ( 2.6%) | 29 ( 2.2%) | 43 ( 3.2%) |
| Varicella | 228 (17.0%) | 467 (34.8%) | 268 (20.0%) | 185 (13.8%) | 87 ( 6.5%) | 37 ( 2.8%) | 28 ( 2.1%) | 41 ( 3.1%) |
| Tdap | 302 (22.5%) | 525 (39.1%) | 248 (18.5%) | 131 ( 9.8%) | 57 ( 4.3%) | 18 ( 1.3%) | 20 ( 1.5%) | 40 ( 3.0%) |
| HPV | 68 ( 5.1%) | 158 (11.8%) | 169 (12.6%) | 255 (19.0%) | 244 (18.2%) | 134 (10.0%) | 171 (12.8%) | 142 (10.6%) |
| MenACWY | 224 (16.7%) | 409 (30.5%) | 272 (20.3%) | 185 (13.8%) | 112 ( 8.4%) | 33 ( 2.5%) | 43 ( 3.2%) | 63 ( 4.7%) |
| Influenza | 57 ( 4.3%) | 90 ( 6.7%) | 96 ( 7.2%) | 163 (12.2%) | 233 (17.4%) | 184 (13.7%) | 296 (22.1%) | 222 (16.6%) |
| COVID-19 | 54 ( 4.0%) | 46 ( 3.4%) | 52 ( 3.9%) | 69 ( 5.1%) | 107 ( 8.0%) | 96 ( 7.2%) | 225 (16.8%) | 692 (51.6%) |
| Adult Vaccines* |  |  |  |  |  |  |  |  |
| Shingles | 41 ( 4.7%) | 87 (10.0%) | 112 (12.8%) | 162 (18.5%) | 186 (21.3%) | 80 ( 9.2%) | 129 (14.8%) | 77 ( 8.8%) |
| Pneumococcal | 61 ( 7.0%) | 123 (14.1%) | 121 (13.8%) | 173 (19.8%) | 150 (17.2%) | 75 ( 8.6%) | 106 (12.1%) | 65 ( 7.4%) |
| RSV | 55 ( 6.3%) | 84 ( 9.6%) | 80 ( 9.2%) | 136 (15.6%) | 126 (14.4%) | 74 ( 8.5%) | 164 (18.8%) | 155 (17.7%) |
| Tdap | 185 (15.9%) | 277 (23.8%) | 245 (21.1%) | 166 (14.3%) | 119 (10.2%) | 48 ( 4.1%) | 65 ( 5.6%) | 57 ( 4.9%) |
| Influenza | 41 ( 3.5%) | 68 ( 5.9%) | 89 ( 7.7%) | 177 (15.2%) | 219 (18.8%) | 153 (13.2%) | 247 (21.3%) | 168 (14.5%) |
| COVID-19 | 25 ( 2.2%) | 45 ( 3.9%) | 38 ( 3.3%) | 74 ( 6.4%) | 133 (11.4%) | 112 ( 9.6%) | 222 (19.1%) | 513 (44.1%) |

\*Respondents were asked whether they provided care for children, young adults, and/or older adults, and were only presented survey items pertaining to vaccines relevant for their patient populations

**Supplemental Table 8.**
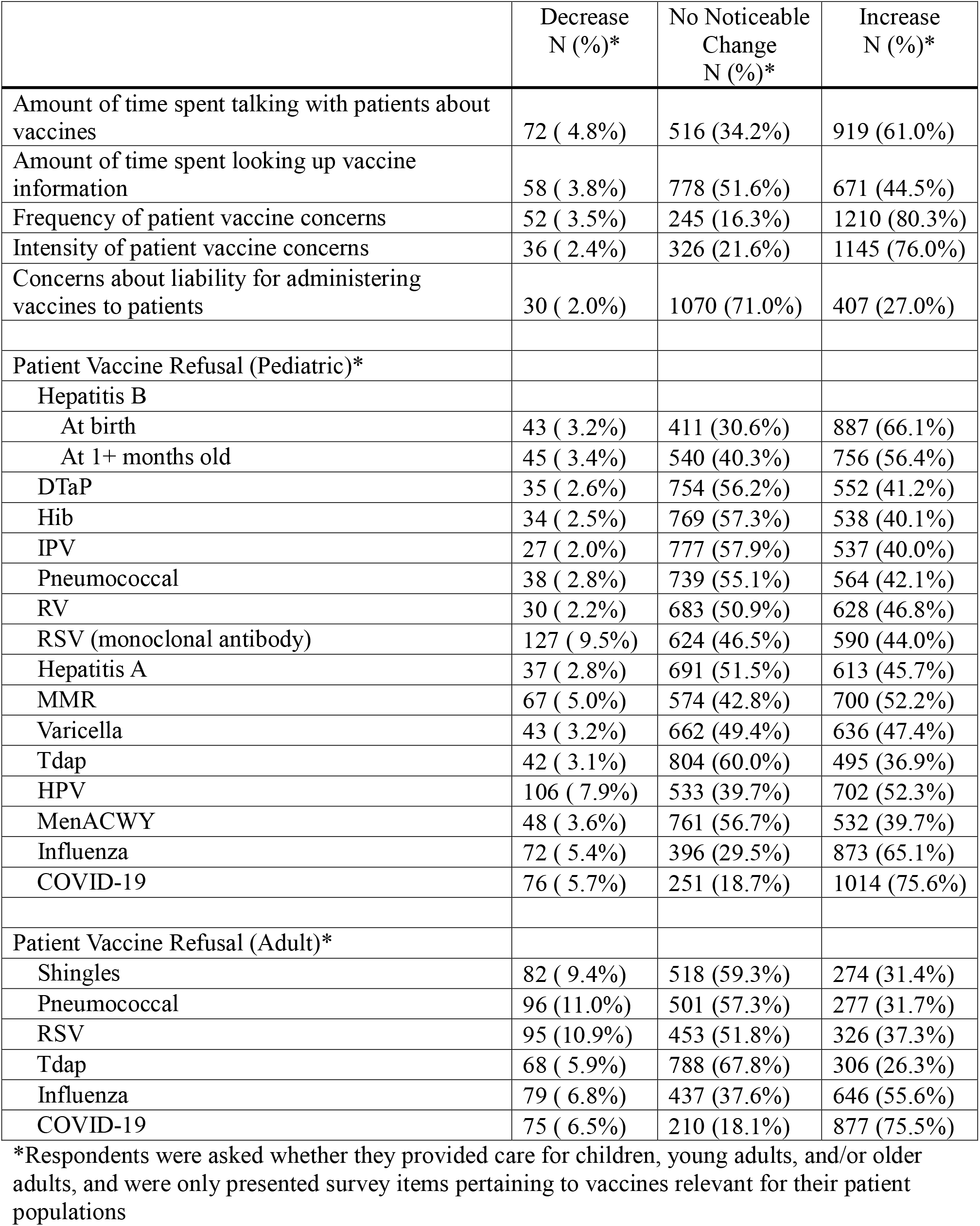
Reported Changes in Vaccine Concerns and Refusal from June 2025 to July 2026.

**Supplemental Table 9.** Proportion of Clinicians Recommending Specific Vaccines for Patients, Stratified by Trust in Vaccine Recommendations from CDC.

|  | Total<br>N (%)* | Low Trust<br>N (%)* | High Trust<br>N (%)* | p-value |
|---|---|---|---|---|
| All | N=1,505 | N=801 | N=704 |  |
| Recommendations for Specific Vaccines |  |  |  |  |
| Hepatitis B |  |  |  |  |
| At birth | 1,073 (89.3%) | 622 (91.3%) | 451 (86.7%) | **0.01** |
| At 1-2 months old | 1,148 (92.1%) | 657 (93.6%) | 491 (90.1%) | **0.02** |
| At 6-18 months old | 1,173 (92.4%) | 665 (93.8%) | 508 (90.7%) | **0.04** |
| Routine childhood vaccines (e.g., MMR, DTaP)<sup>2</sup> | 644 (97.3%) | 353 (97.2%) | 291 (97.3%) | 0.95 |
| DTaP<sup>1</sup> | 645 (97.7%) | 356 (96.7%) | 289 (99.0%) | 0.056 |
| Hib<sup>1</sup> | 639 (97.0%) | 356 (96.7%) | 283 (97.3%) | 0.70 |
| IPV<sup>1</sup> | 637 (96.8%) | 357 (97.3%) | 280 (96.2%) | 0.44 |
| Pneumococcal<sup>1</sup> | 640 (96.8%) | 359 (97.3%) | 281 (96.2%) | 0.44 |
| RV<sup>1</sup> | 602 (92.0%) | 348 (94.6%) | 254 (88.8%) | **<0.01** |
| RSV (monoclonal antibody)<sup>1</sup> | 598 (91.4%) | 342 (93.4%) | 256 (88.9%) | **0.04** |
| Hepatitis A<sup>1</sup> | 597 (90.0%) | 345 (93.2%) | 252 (86.0%) | **<0.01** |
| MMR<sup>1</sup> | 652 (98.0%) | 361 (97.3%) | 291 (99.0%) | 0.12 |
| Varicella<sup>1</sup> | 646 (97.3%) | 359 (97.0%) | 287 (97.6%) | 0.64 |
| MMRV<sup>1</sup> | 612 (92.2%) | 342 (92.2%) | 270 (92.2%) | 0.99 |
| Routine adolescent vaccines |  |  |  |  |
| Tdap<sup>1</sup> | 655 (97.9%) | 365 (97.6%) | 290 (98.3%) | 0.52 |
| MenACWY<sup>1</sup> | 648 (96.9%) | 362 (96.8%) | 286 (96.9%) | 0.91 |
| HPV (all ages)<sup>2</sup> | 648 (88.5%) | 346 (88.7%) | 302 (88.3%) | 0.86 |
| 9-10 years old<sup>1</sup> | 400 (60.3%) | 223 (60.3%) | 177 (60.4%) | 0.97 |
| 11-12 years old<sup>1</sup> | 582 (87.4%) | 334 (90.0%) | 248 (84.1%) | **0.02** |
| 13-15 years old<sup>1</sup> | 629 (94.0%) | 353 (94.6%) | 276 (93.2%) | 0.45 |
| 16-17 years old<sup>1</sup> | 628 (93.6%) | 350 (93.6%) | 278 (93.6%) | 0.99 |
| 18-26 years old<sup>1</sup> | 510 (89.8%) | 256 (91.4%) | 254 (88.2%) | 0.20 |
| 27-45 years old<sup>1</sup> | 301 (60.9%) | 140 (61.9%) | 161 (60.1%) | 0.67 |
| Respiratory vaccines |  |  |  |  |
| Influenza (all ages)<sup>2</sup> | 677 (89.8%) | 349 (87.3%) | 328 (92.7%) | **0.01** |
| 6-23 months old<sup>1</sup> | 596 (92.0%) | 339 (93.4%) | 257 (90.2%) | 0.14 |
| 2-4 years old<sup>1</sup> | 602 (92.5%) | 345 (94.0%) | 257 (90.5%) | 0.09 |
| 5-11 years old<sup>1</sup> | 600 (90.8%) | 338 (91.4%) | 262 (90.0%) | 0.56 |
| 12-15 years old<sup>1</sup> | 598 (89.4%) | 340 (91.2%) | 258 (87.2%) | 0.10 |
| 16-17 years old<sup>1</sup> | 596 (88.6%) | 339 (90.4%) | 257 (86.2%) | 0.09 |
| 18-49 years old<sup>1</sup> | 516 (91.3%) | 256 (92.8%) | 260 (90.0%) | 0.24 |
| 50-64 years old<sup>1</sup> | 422 (97.2%) | 180 (95.7%) | 242 (98.4%) | 0.10 |
| 65 and above<sup>1</sup> | 423 (98.1%) | 181 (97.3%) | 242 (98.8%) | 0.26 |
| Pneumococcal (all ages)<sup>2</sup> | 723 (96.8%) | 381 (96.2%) | 342 (97.4%) | 0.34 |
| 50-64 years old<sup>1</sup> | 387 (89.6%) | 169 (90.9%) | 218 (88.6%) | 0.45 |
| 65 and above<sup>1</sup> | 424 (98.1%) | 181 (97.3%) | 243 (98.8%) | 0.26 |
| RSV |  |  |  |  |
| 50-59 years old (with risk factor) | 603 (69.6%) | 268 (70.3%) | 335 (68.9%) | 0.65 |
| 60-74 years old (with risk factor) | 754 (87.0%) | 329 (86.1%) | 425 (87.6%) | 0.51 |
| 75 and above | 776 (90.1%) | 334 (88.1%) | 442 (91.7%) | 0.081 |
| COVID-19 |  |  |  |  |
| 6-23 months old | 546 (42.6%) | 330 (46.2%) | 216 (38.1%) | **<0.01** |
| 2-4 years old | 519 (40.3%) | 307 (42.6%) | 212 (37.5%) | 0.06 |
| 5-11 years old | 528 (40.3%) | 308 (42.2%) | 220 (37.9%) | 0.11 |
| 12-15 years old | 540 (40.7%) | 310 (42.2%) | 230 (38.9%) | 0.22 |
| 16-17 years old | 549 (41.1%) | 314 (42.5%) | 235 (39.3%) | 0.24 |
| 18-24 years old | 512 (45.0%) | 262 (46.3%) | 250 (43.7%) | 0.38 |
| 25-49 years old | 504 (50.2%) | 241 (51.7%) | 263 (48.9%) | 0.37 |
| 50-64 years old | 584 (67.2%) | 271 (70.9%) | 313 (64.3%) | **0.04** |
| 65 and above | 668 (76.9%) | 302 (79.1%) | 366 (75.2%) | 0.18 |
| Other routine adult vaccines |  |  |  |  |
| Shingles (all ages)<sup>2</sup> | 412 (94.5%) | 182 (93.3%) | 230 (95.4%) | 0.34 |
| 50-59 years old<sup>1</sup> | 400 (92.4%) | 174 (93.0%) | 226 (91.9%) | 0.65 |
| 60-74 years old<sup>1</sup> | 420 (96.8%) | 179 (95.2%) | 241 (98.0%) | 0.11 |
| 75 and above<sup>1</sup> | 408 (95.1%) | 173 (93.0%) | 235 (96.7%) | 0.08 |

\*Respondents were asked whether they provided care for children, young adults, and/or older adults, and were only presented survey items pertaining to vaccines relevant for their patient populations; half of respondents were randomized to receive items asking about their recommendations for specific vaccines and age groups<sup>1</sup>, while the other half received items asking about their vaccine recommendations more generally<sup>2</sup> (to facilitate direct comparison with a previous survey);<sup>19</sup> numbers and percentages presented in table refer to clinicians recommending or strongly recommending the vaccine in each row.

**Supplemental Table 10.** Reported Increases in Vaccine Concerns and Refusal from June 2025 to July 2026, Stratified by Trust in Vaccine Recommendations from CDC.

|  | Total<br>N (%)* | Low Trust<br>N (%)* | High Trust<br>N (%)* | p-value |
|---|---|---|---|---|
| All | N=1,505 | N=801 | N=704 |  |
| Amount of time spent talking with patients about vaccines | 917 (60.9%) | 541 (67.5%) | 376 (53.4%) | **<0.01** |
| Amount of time spent looking up vaccine information | 670 (44.5%) | 350 (43.7%) | 320 (45.5%) | 0.49 |
| Frequency of patient vaccine concerns | 1,208 (80.3%) | 686 (85.6%) | 522 (74.1%) | **<0.01** |
| Intensity of patient vaccine concerns | 1,143 (75.9%) | 646 (80.6%) | 497 (70.6%) | **<0.01** |
| Concerns about liability for administering vaccines | 407 (27.0%) | 231 (28.8%) | 176 (25.0%) | 0.09 |
| Patient Vaccine Refusal (Pediatric) |  |  |  |  |
| Hepatitis B |  |  |  |  |
| At birth | 886 (66.2%) | 552 (74.7%) | 334 (55.7%) | **<0.01** |
| At 1+ months old | 755 (56.4%) | 465 (62.9%) | 290 (48.3%) | **<0.01** |
| DTaP | 551 (41.2%) | 334 (45.2%) | 217 (36.2%) | **<0.01** |
| Hib | 537 (40.1%) | 333 (45.1%) | 204 (34.0%) | **<0.01** |
| IPV | 536 (40.0%) | 323 (43.7%) | 213 (35.5%) | **<0.01** |
| Pneumococcal | 563 (42.0%) | 346 (46.8%) | 217 (36.2%) | **<0.01** |
| RV | 627 (46.8%) | 383 (51.8%) | 244 (40.7%) | **<0.01** |
| RSV (monoclonal antibody) | 589 (44.0%) | 355 (48.0%) | 234 (39.0%) | **<0.01** |
| Hepatitis A | 612 (45.7%) | 380 (51.4%) | 232 (38.7%) | **<0.01** |
| MMR | 699 (52.2%) | 427 (57.8%) | 272 (45.3%) | **<0.01** |
| Varicella | 635 (47.4%) | 389 (52.6%) | 246 (41.0%) | **<0.01** |
| Tdap | 494 (36.9%) | 300 (40.6%) | 194 (32.3%) | **<0.01** |
| HPV | 700 (52.3%) | 418 (56.6%) | 282 (47.0%) | **<0.01** |
| MenACWY | 531 (39.7%) | 325 (44.0%) | 206 (34.3%) | **<0.01** |
| Influenza | 871 (65.0%) | 518 (70.1%) | 353 (58.8%) | **<0.01** |
| COVID-19 | 1,012 (75.6%) | 574 (77.7%) | 438 (73.0%) | **0.05** |
| Patient Vaccine Refusal (Adult) |  |  |  |  |
| Shingles | 274 (31.4%) | 133 (34.6%) | 141 (28.9%) | 0.07 |
| Pneumococcal | 277 (31.8%) | 132 (34.4%) | 145 (29.7%) | 0.14 |
| RSV | 324 (37.2%) | 152 (39.6%) | 172 (35.2%) | 0.19 |
| Tdap | 306 (26.4%) | 171 (29.7%) | 135 (23.1%) | **0.01** |
| Influenza | 644 (55.5%) | 349 (60.7%) | 295 (50.4%) | **<0.01** |
| COVID-19 | 875 (75.4%) | 454 (79.0%) | 421 (72.0%) | **<0.01** |

\*Respondents were asked whether they provided care for children, young adults, and/or older adults, and were only presented survey items pertaining to vaccines relevant for their patient populations

**Supplemental Figure 1.**
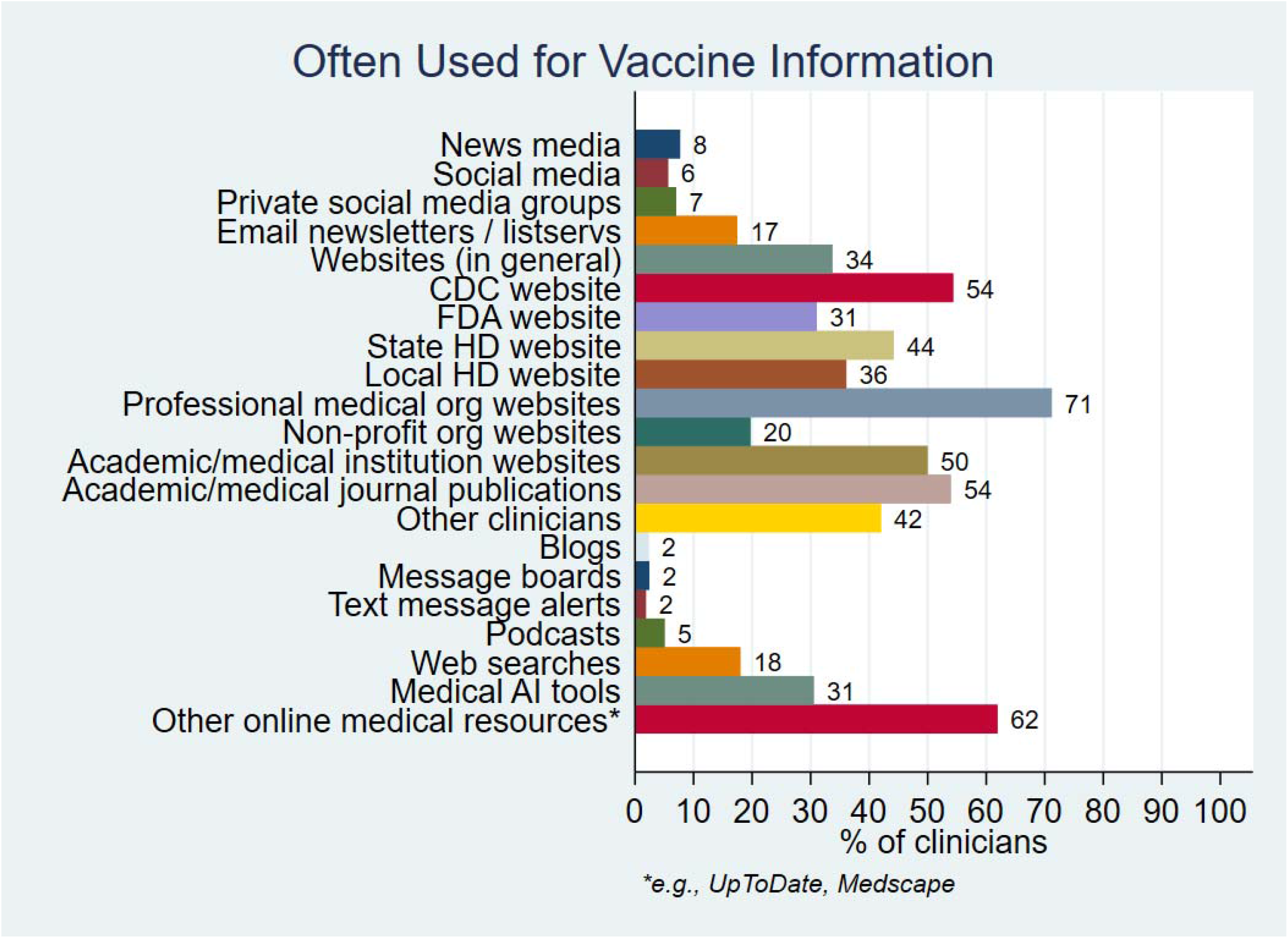
Sources of Vaccine Information Used by Clinicians.

**Supplemental Figure 2.**
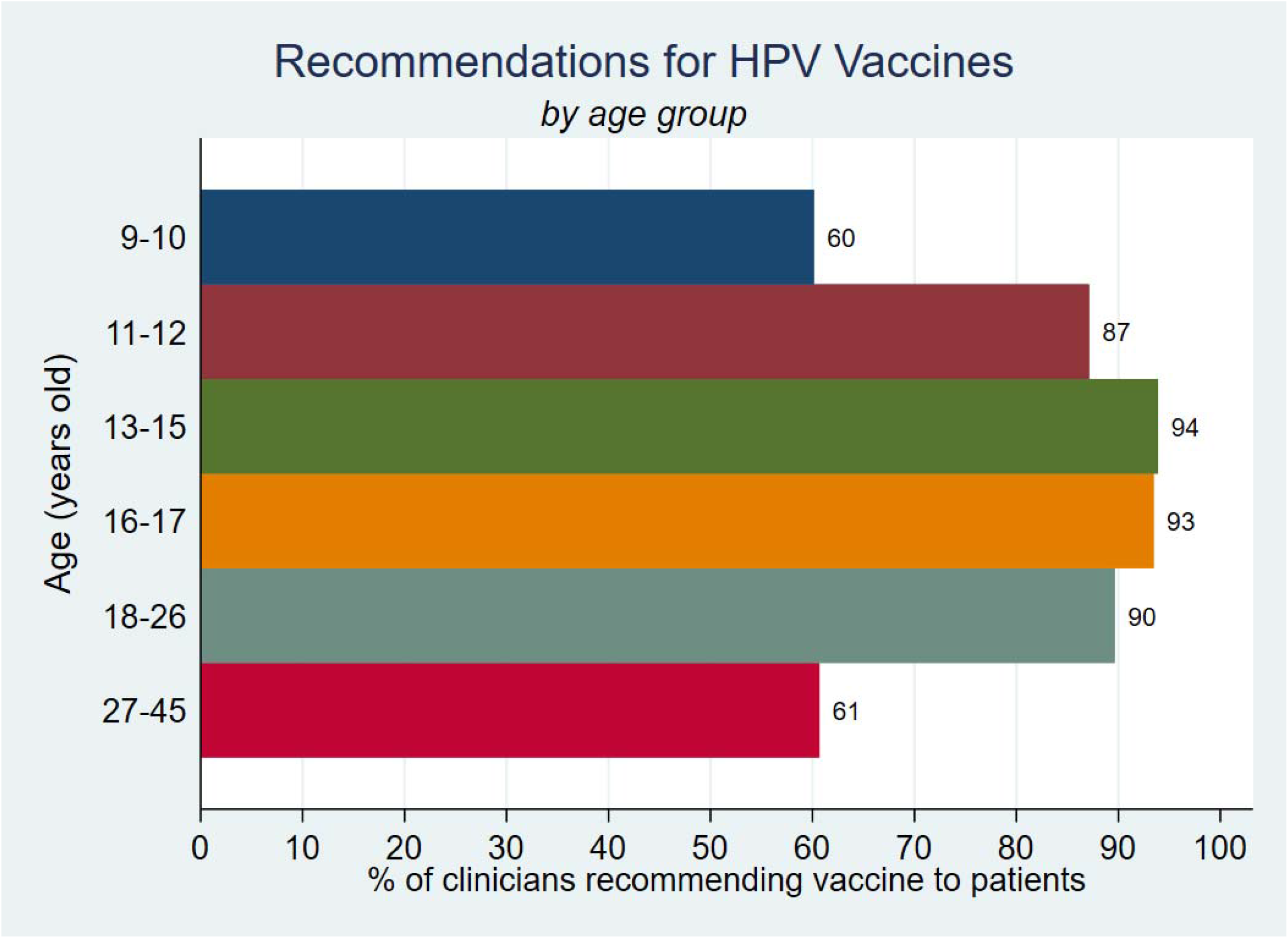
Strength of HPV Vaccine Recommendations, Stratified by Age Group.

**Supplemental Figure 3.**
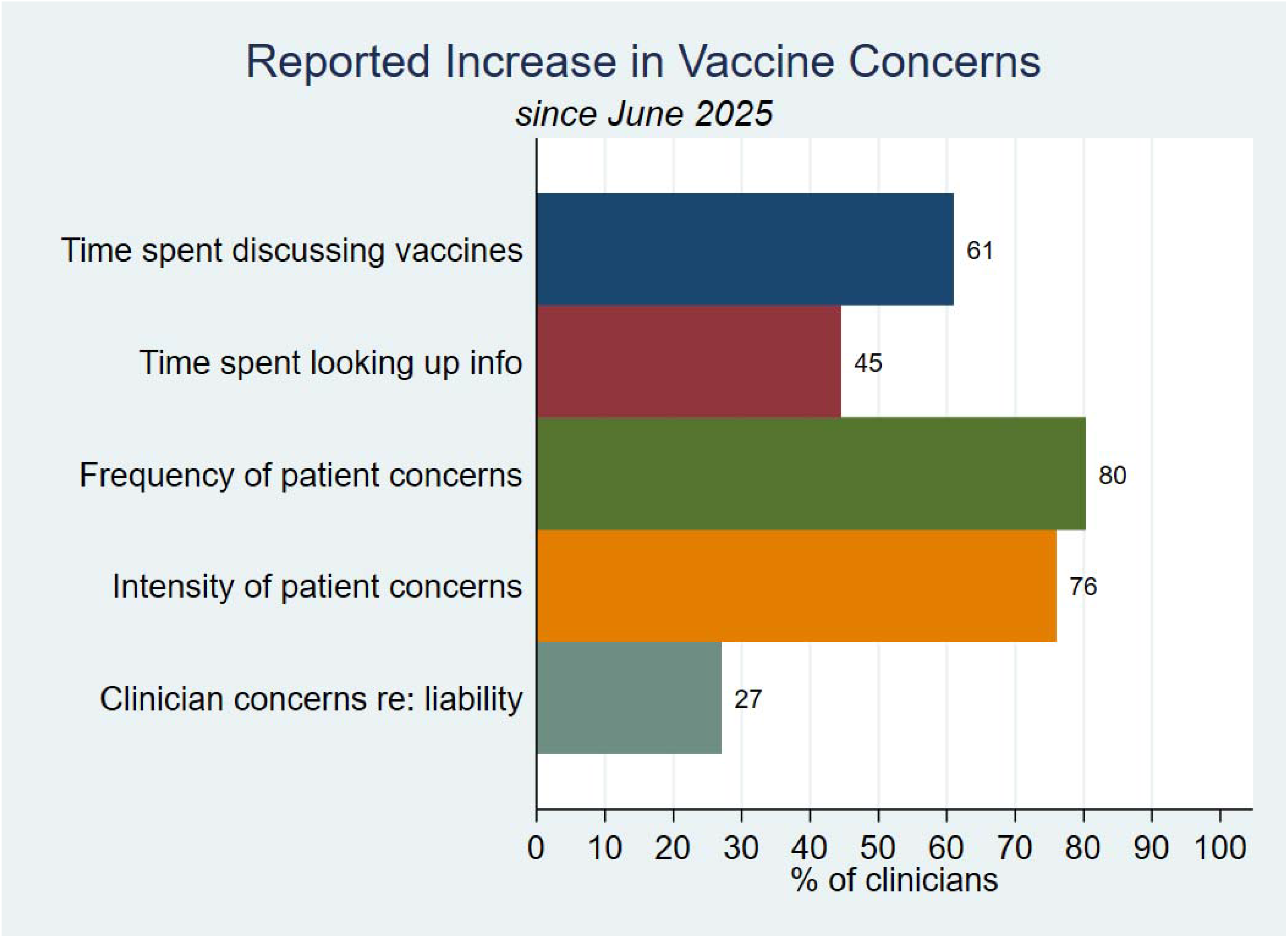
Reported Changes in Clinician Vaccination Experiences from June 2025 to July 2026.

**Supplemental Figure 4.**
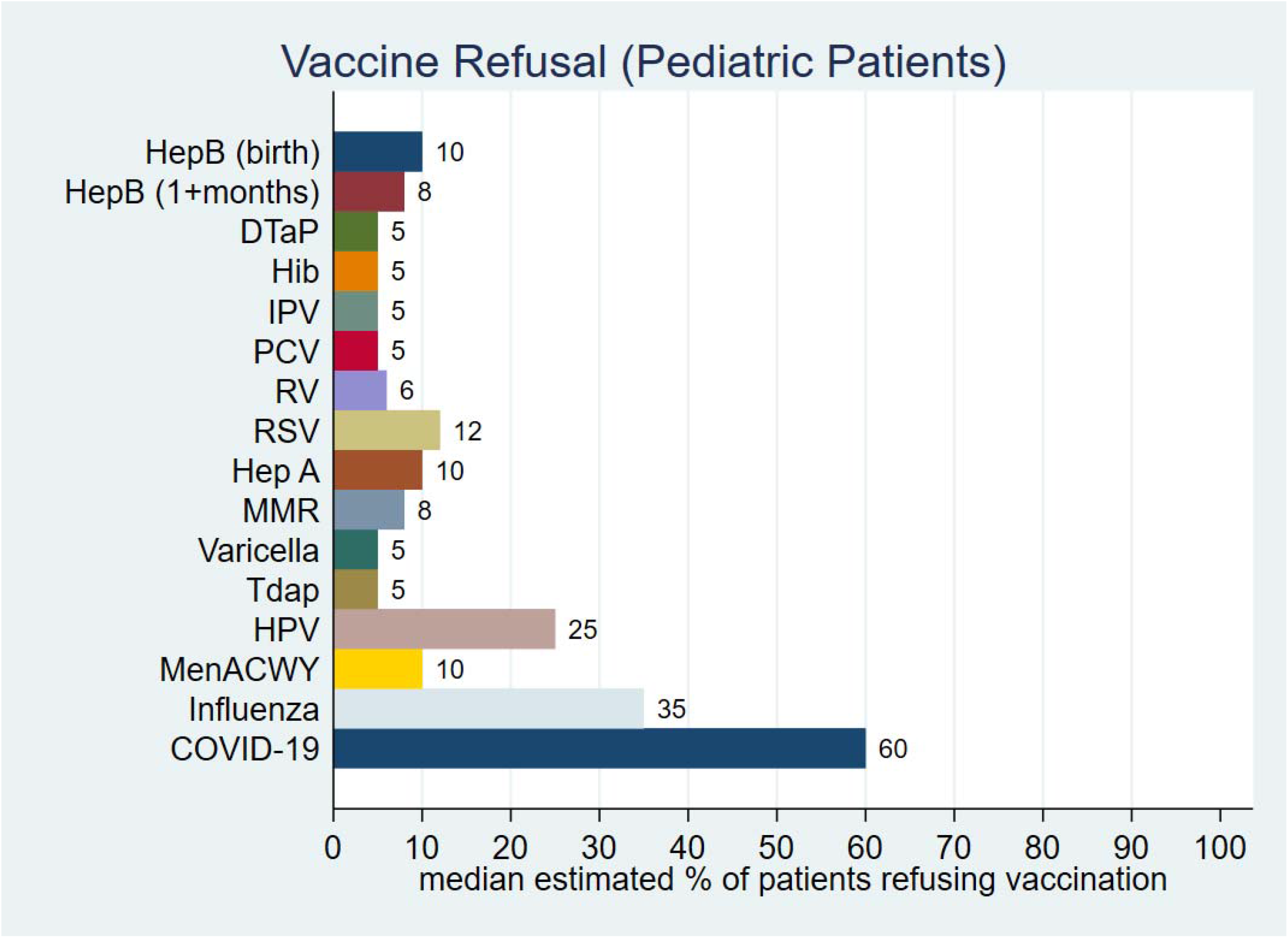
Median Estimated Pediatric Patient Vaccine Refusal Rates from June 2025 to July 2026.

**Supplemental Figure 5.**
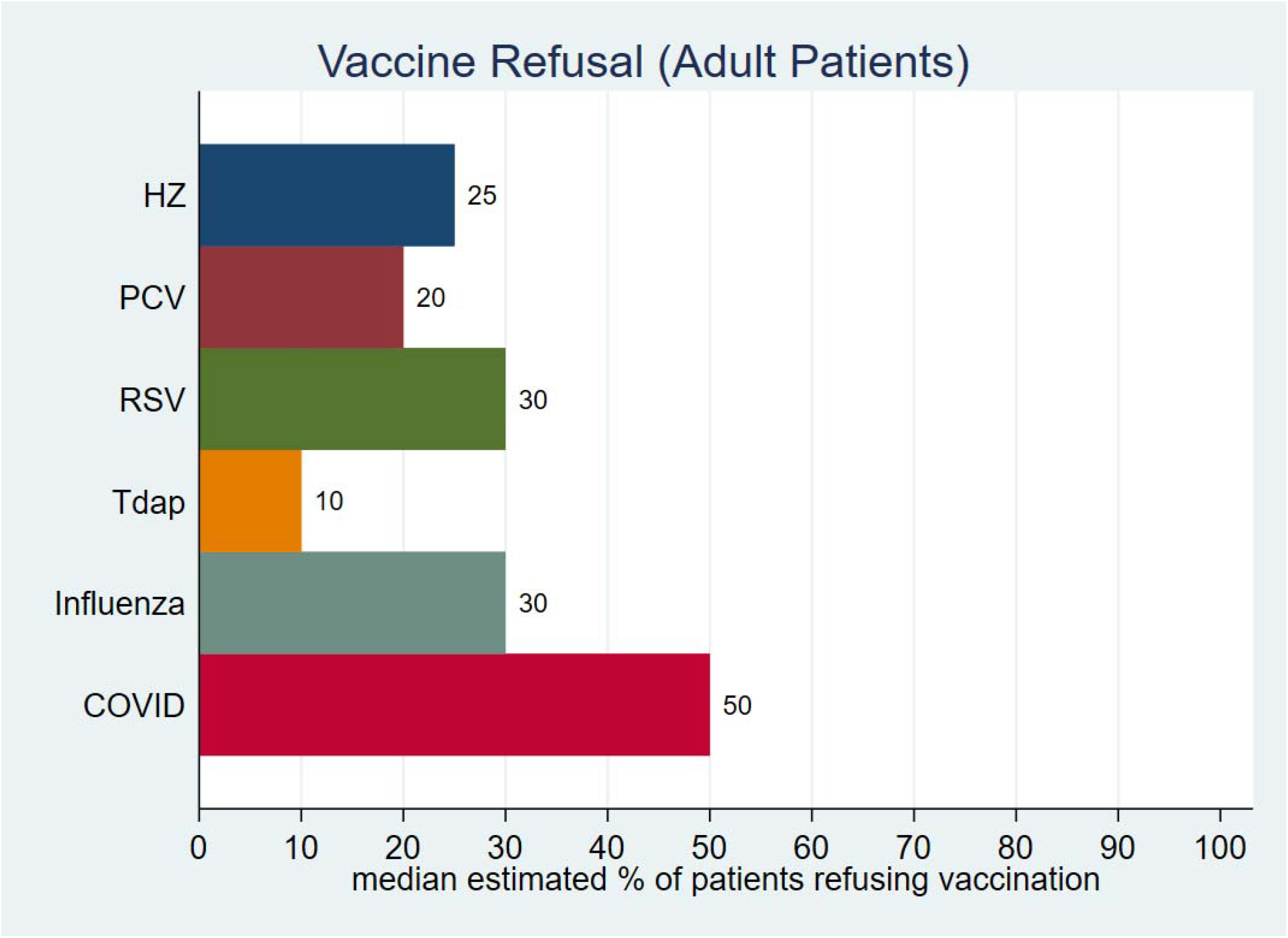
Median Estimated Adult Patient Vaccine Refusal Rates from June 2025 to July 2026.

**Supplemental Material 1. Survey Instrument**

Note: The questions in black text are excerpts from the most recent survey instrument we used for clinicians (in January 2023) to enable accurate comparison between timepoints. New questions that were not included in the previous survey (along with any updates to existing questions) are indicated by red text.

## Introduction

This survey’s purpose is to learn how changes made to the CDC’s immunization schedule over the past year have affected clinicians’ practice and experience with patients. The survey is voluntary, anonymous, and should take about 10 minutes to complete.

S1: What is your current medical profession?

a. Physician Go to Question S1a; skip Question S1b
b. Physician Assistant Go to Question S1b.
c. Nurse Practitioner Go to Question S1b.
d. Nurse (Registered Nurse or Licensed Practical Nurse) Go to Question S1b.
e. None/Other (Specify): _________TERMINATE

S1a: [PN: Show if S1=a] Which of the following best describes your medical specialty?

a. Primary Care: Internal Medicine TERMINATE
b. Primary Care: Family Practice
c. Primary Care: General Pediatrics
d. General Surgery TERMINATE
e. Dermatology TERMINATE
f. Rheumatology TERMINATE
g. Emergency Medicine TERMINATE
h. Other (Specify): ______________ TERMINATE

S1b: [PN: Show if S1 = b, c, OR d ] Which of the following best describes the specialty setting in which you currently practice?

a. Primary Care: Internal Medicine
b. Primary Care: Family Practice
c. Primary Care: General Pediatrics
d. Other (Specify): ______________ TERMINATE

S2. Do you provide direct patient care (the provision of health care services directly to individuals, i.e. administration of medication, patient counseling, etc.)

a. Yes
b. No Thank you for your time, this survey is intended for those who provide direct patient care.

S3. Does your practice currently administer vaccines?

a. Yes
b. No Thank you for your time, this survey is intended for those who administer vaccines.

S4. Which of the following best describes your practice location:

a. Urban [more than 50,000 people]
b. Suburban [within metropolitan areas (not rural) and outside the central cities (not core)]
c. Rural [up to 50,000 people]

S5. In which U.S. state is your practice located? Please specify: __ [PN: State Drop Down] ___________

### Section One: General Practice Background

*First, we’d like to understand the nature of your current practice setting*.

1. Who do you provide care for? (check all that apply)

a. Children (<18 years of age) ◊ [PN: Show questions pertaining to children, denoted with **]
b. Young Adults (18-49 years of age) ◊ [PN: Show questions pertaining to young adults, denoted with *]
c. Older Adults (≥50 years of age) ◊ [PN: Show questions pertaining to older adults, denoted with ***]
2. On average, how many patients do you see per day? ____________
3. How many years have you been in practice? ____________

*Instructions: The following questions pertain to your primary site of outpatient practice. If you practice at more than one outpatient site, please respond to these questions in terms of the site at which you spend the most time*.

4. Which of the following best describes your practice setting:

a. Private, independent practice
b. Practice network/ HMO
c. Hospital or medical center
d. Community health center/Federally Qualified Health Center (FQHC)
e. Other

1) If you chose ‘Other’, please specify your practice type: ________________________
5. In your primary outpatient practice, roughly what percentages of your patients are in the following groups? (Please approximate; groups do not have to sum up to 100%) ____________ [PN: display automatic total]

5.1 Private Insurance
5.2 Medicaid or your state’s Children’s Health Insurance Program (CHIP)
5.3 Medicare
5.4 Uninsured
6. In your primary service location, roughly what percentages of your patients are in the following racial/ethnic groups? (Please approximate; groups do not have to sum up to 100%) ____________ [PN: display automatic total]

6.1 Hispanic or Latino
6.2 Black or African American
6.3 Asian
6.4 White
6.5 Other
7. Does your practice use Electronic Health Records (EHR)?

a. Yes

1) If so, which one? _____
b. No
8. Does your practice participate in the Vaccines for Children (VFC) program?** [Base: Ask If Q1=a]

a. Yes
b. No, and we never have
c. No, but we did previously
d. Unsure
9. During which of the following years did your practice provide seasonal influenza vaccinations to eligible patients? (check all that apply)

a. 2023-2024
b. 2024-2025
c. 2025-2026
d. Did not administer the influenza vaccine in any of these years

### Section Two: Vaccine Recommendations for Patients

*The purpose of this section is to learn if/how you currently recommend vaccines for your patients*.

10. How strongly do you recommend **hepatitis B vaccination** for your pediatric patients (whose mothers test negative for hepatitis B virus) in these age groups?** [Base: Ask If Q1=a]

a. 1 (very strongly recommend *against* receiving it)
b. 2
c. 3
d. 4
e. 5 (very strongly recommend *for* receiving it)
f. I do not see patients in this age group

1) At birth
2) 1-2 months old
3) 6-18 months old

[PN: Randomly assign respondents to receive EITHER the next five questions (Q11-15, in black text) OR the following five questions (Q16-20, in red text)]

11. How strongly do you recommend **routine childhood vaccines** (e.g., MMR, DTaP) for your patients?** [Base: Ask If Q1=a]

a. 1 (very strongly recommend *against* receiving them)
b. 2
c. 3
d. 4
e. 5 (very strongly recommend *for* receiving them)
f. I do not see patients in this age group
12. How strongly do you recommend the **HPV vaccine** for your patients?*/** [Base: Ask If Q1=a AND/OR Q1=b]

a. 1 (very strongly recommend *against* receiving it)
b. 2
c. 3
d. 4
e. 5 (very strongly recommend *for* receiving it)
f. I do not see patients in this age group
13. How strongly do you recommend **influenza vaccine** for your patients?

a. 1 (very strongly recommend *against* receiving it)
b. 2
c. 3
d. 4
e. 5 (very strongly recommend *for* receiving it)
14. How strongly do you recommend **pneumococcal vaccine** for your patients who are eligible to receive it?

a. 1 (very strongly recommend *against* receiving it)
b. 2
c. 3
d. 4
e. 5 (very strongly recommend *for* receiving it)
f. I do not see patients in this age group
15. How strongly do you recommend **shingles vaccine** for your patients who are eligible to receive it?*** [Base: Ask If Q1=c]

a. 1 (very strongly recommend *against* receiving it)
b. 2
c. 3
d. 4
e. 5 (very strongly recommend *for* receiving it)
f. I do not see patients in this age group

[PN: Only those who were randomly assigned NOT to receive the above five questions (Q11-15, in black text) will be assigned the following five questions (Q16-20, in red text)]

16. How strongly do you recommend the following vaccines for your pediatric patients who are eligible to receive them?** [Base: Ask If Q1=a]

a. 1 (very strongly recommend *against* receiving them)
b. 2
c. 3
d. 4
e. 5 (very strongly recommend *for* receiving them)
f. I do not see patients in this age group

1) Diphtheria, tetanus, & acellular pertussis (DTaP)
2) *Haemophilus influenzae* type b (Hib)
3) Inactivated poliovirus (IPV)
4) Pneumococcal conjugate (PCV)
5) Rotavirus (RV)
6) Respiratory syncytial virus (RSV)
7) Hepatitis A
8) Measles, mumps, rubella (MMR)
9) Varicella
10) Measles, mumps, rubella, varicella combination vaccine (MMRV)
11) Tetanus, diphtheria, & acellular pertussis (Tdap)
12) Meningococcal (MenACWY)
17. How strongly do you recommend **HPV vaccination** for your eligible patients in these age groups?

a. 1 (very strongly recommend *against* receiving it)
b. 2
c. 3
d. 4
e. 5 (very strongly recommend *for* receiving it)
f. I do not see patients in this age group

1) 9-10 years old** [Base: Ask If Q1=a]
2) 11-12 years old** [Base: Ask If Q1=a]
3) 13-15 years old** [Base: Ask If Q1=a]
4) 16-17 years old** [Base: Ask If Q1=a]
5) 18-26 years old* [Base: Ask If Q1=b]
6) 27-45 years old* [Base: Ask If Q1=b]
18. How strongly do you recommend seasonal **influenza vaccination** for your patients in these age groups?

a. 1 (very strongly recommend *against* receiving it)
b. 2
c. 3
d. 4
e. 5 (very strongly recommend *for* receiving it)
f. I do not see patients in this age group

1) 6-23 months old** [Base: Ask If Q1=a]
2) 2-4 years old** [Base: Ask If Q1=a]
3) 5-11 years old** [Base: Ask If Q1=a]
4) 12-15 years old** [Base: Ask If Q1=a]
5) 16-17 years old** [Base: Ask If Q1=a]
6) 18-49 years old* [Base: Ask If Q1=b]
7) 50-64 years old*** [Base: Ask If Q1=c]
8) 65 and above*** [Base: Ask If Q1=c]
19. How strongly do you recommend **pneumococcal vaccination** for your adult patients in these age groups?*** [Base: Ask If Q1=c]

a. 1 (very strongly recommend *against* receiving it)
b. 2
c. 3
d. 4
e. 5 (very strongly recommend *for* receiving it)
f. I do not see patients in this age group

1) 50-64 years old
2) 65 and above
20. How strongly do you recommend **shingles vaccination** for your adult patients in these age groups?*** [Base: Ask If Q1=c]

a. 1 (very strongly recommend *against* receiving it)
b. 2
c. 3
d. 4
e. 5 (very strongly recommend *for* receiving it)
f. I do not see patients in this age group

1) 50-59 years old
2) 60-74 years old
3) 75 and above
21. How strongly do you recommend **respiratory syncytial virus (RSV) vaccination** for your adult patients in these age groups?*** [Base: Ask If Q1=c]

a. 1 (very strongly recommend *against* receiving it)
b. 2
c. 3
d. 4
e. 5 (very strongly recommend *for* receiving it)
f. I do not see patients in this age group

1) 50-59 years old (with risk factors)
2) 60-74 years old (with risk factors)
3) 75 and above
22. How strongly do you recommend **COVID-19 vaccination** for your patients in these age groups?

a. 1 (very strongly recommend *against* receiving it)
b. 2
c. 3
d. 4
e. 5 (very strongly recommend *for* receiving it)
f. I do not see patients in this age group

1) 6-23 months old** [Base: Ask If Q1=a]
2) 2-4 years old** [Base: Ask If Q1=a]
3) 5-11 years old** [Base: Ask If Q1=a]
4) 12-15 years old** [Base: Ask If Q1=a]
5) 16-17 years old** [Base: Ask If Q1=a]
6) 18-24 years old* [Base: Ask If Q1=b]
7) 25-49 years old* [Base: Ask If Q1=b]
8) 50-64 years old*** [Base: Ask If Q1=c]
9) 65 and above*** [Base: Ask If Q1=c]

### Section Three: Vaccine Discussions with Patientsh

*The purpose of this section is to learn how you discuss vaccines with your patients*.

23. How much time *each week* (on average) do you spend talking with patients about vaccines? ___ [PN: include dropdown menu to choose between minutes and hours]
24. How much time *during each clinical encounter in which a vaccine is due* (on average) do you spend talking with patients about vaccines? ___ [PN: include dropdown menu to choose between minutes and hours]
25. Over the past year, how has the amount of time you spend talking with patients about vaccines changed?

a. It increased
b. It decreased
c. No noticeable change
26. How much time *each week* (on average) do you spend looking up vaccine information (e.g., recommendations, data on safety/effectiveness)? ___ [PN: include dropdown menu to choose between minutes and hours]
27. Over the past year, how has the amount of time you spend looking up vaccine information changed?

a. It increased
b. It decreased
c. No noticeable change
28. Roughly what percentage of your *pediatric* patients’ parents have concerns about vaccines?** ____________ [Base: Ask If Q1=a]
29. Roughly what percentage of your *adult* patients have concerns about vaccines?*/*** ____________ [Base: Ask If Q1=b AND/OR Q1=c]
30. Over the past year, how has the *frequency* of vaccine concerns among your patients changed?

a. It *increased* (more patients have vaccine concerns now)
b. It *decreased* (fewer patients have vaccine concerns now)
c. No noticeable change
31. Over the past year, how has the *intensity* of vaccine concerns among your patients changed?

a. It *increased* (patients with vaccine concerns became more concerned)
b. It *decreased* (patients with vaccine concerns became less concerned)
c. No noticeable change
32. What are the most common vaccine concerns among your patients right now? _______________
33. Over the past year:

33_NUM. Roughly what percentage of your *pediatric* patients eligible to receive these vaccines missed at least one recommended dose *due to parent refusal*?** ___ [Base: Ask If Q1=a]
33_Radio. How has the frequency of vaccine refusals among your *pediatric* patients changed?** [Base: Ask If Q1=a]

a. It *increased* (more refused to vaccinate)
b. It *decreased* (fewer refused to vaccinate)
c. No noticeable change

1) Hepatitis B (birth dose)
2) Hepatitis B (after at least 1 month of age)
3) Diphtheria, tetanus, & acellular pertussis (DTaP)
4) *Haemophilus influenzae* type b (Hib)
5) Inactivated poliovirus (IPV)
6) Pneumococcal conjugate (PCV)
7) Rotavirus (RV)
8) Respiratory syncytial virus (RSV)
9) Hepatitis A
10) Measles, mumps, rubella (MMR)
11) Varicella
12) Tetanus, diphtheria, & acellular pertussis (Tdap)
13) Human papillomavirus (HPV)
14) Meningococcal (MenACWY)
15) Influenza
16) COVID-19

34. Over the past year:

34_NUM. Roughly what percentage of your *adult* patients eligible to receive these vaccines *refused* to receive at least one recommended dose? ___
34_Radio. How has the frequency of vaccine refusals among your *adult* patients changed?

a. It *increased* (more refused to vaccinate)
b. It *decreased* (fewer refused to vaccinate)
c. No noticeable change

1) Herpes zoster (shingles)*** [Base: Ask If Q1=c]
2) Pneumococcal*** [Base: Ask If Q1=c]
3) Respiratory syncytial virus (RSV)*** [Base: Ask If Q1=c]
4) Tetanus, diphtheria, & acellular pertussis (Tdap)*/*** [Base: Ask If Q1=b AND/OR Q1=c]
5) Influenza*/*** [Base: Ask If Q1=b AND/OR Q1=c]
6) COVID-19*/*** [Base: Ask If Q1=b AND/OR Q1=c]

35. How well prepared do you feel for conversations with patients about vaccines?

a. 1 (not at all prepared)
b. 2
c. 3
d. 4
e. 5 (very well prepared)
[PN: Randomly assign respondents to receive EITHER the next question (Q36, in black text) OR the following two questions (Q37-38, in red text)]
36. How easy is it for you to stay up to date on vaccine recommendations, contraindications, and vaccine controversies?
1 (not easy at all)
2
3
4
5 (very easy)

[PN: Only those who were randomly assigned NOT to receive the above question (Q36, in black text) will be assigned the following two questions (Q37-38, in red text)]

37. How easy is it for you to stay up to date on vaccine recommendations and contraindications?

a. 1 (not easy at all)
b. 2
c. 3
d. 4
e. 5 (very easy)
38. How easy is it for you to stay up to date on vaccine concerns and controversies?

a. 1 (not easy at all)
b. 2
c. 3
d. 4
e. 5 (very easy)
39. Do patients sometimes ask vaccine questions to which you are unsure of the scientific answer?

a. Yes
b. No
40. Would it be helpful to know a patient’s vaccine intentions and any concerns prior to a clinical encounter when a patient is due a vaccine?

a. Yes
b. No
41. Roughly what percentage of visits *in which you administer a vaccine* does your practice bill for vaccine *administration*?

a. None
b. Less than 25%
c. 25-50%
d. 51-75%
e. More than 75%
f. Unsure
42. Roughly what percentage of visits *in which you administer a vaccine* does your practice bill for vaccine *counseling*?

a. None
b. Less than 25%
c. 25-50%
d. 51-75%
e. More than 75%
f. Unsure
43. Roughly what percentage of visits in which you *discuss* vaccines with patients but do *not* administer a vaccine does your practice bill for vaccine *counseling*?

a. None
b. Less than 25%
c. 25-50%
d. 51-75%
e. More than 75%
f. Unsure
44. Roughly what percentage of visits in which you administer a vaccine do you provide a Vaccine Information Statement (VIS)?

a. None
b. Less than 25%
c. 25-50%
d. 51-75%
e. More than 75%
f. Unsure
45. ow is informed consent for vaccination typically obtained at your practice? (check all that apply)

a. Written/signed consent
b. Verbal/oral consent
c. Digital/electronic consent
d. Implied consent
e. Other (Specify): ______________
46. How concerned are you about liability for administering vaccines to patients?

a. 1 (not at all concerned)
b. 2
c. 3
d. 4
e. 5 (very concerned)
47. Over the past year, how has your level of concern about vaccine-related liability changed?

a. It increased
b. It decreased
c. No noticeable change
48. In what other ways have changes made to the CDC’s immunization schedule over the past year affected your vaccine-related experiences with patients? Please explain in detail. ____

### Section Four: Vaccine Information Sources

*The purpose of this section is to learn how you prefer to receive and share vaccine information*.

49. How much do you *use* each of the following channels to get information about vaccines?

a. 1 (never)
b. 2 (only occasionally)
c. 3 (sometimes)
d. 4 (often)
e. 5 (very often)

News media
Social media
Private social media groups
Email newsletters / listservs
Websites (in general)
Website of the Centers for Disease Control and Prevention (CDC)
Website of the Food and Drug Administration (FDA)
Website of your state public health department
Website of your local public health department
Websites of professional medical organizations
Websites of vaccine-focused non-profit organizations
Websites of academic/medical institutions
Publications in academic/medical journals
Other healthcare providers
Blogs
Message boards
Text message alerts
Podcasts
Web searches (e.g., Google, Bing, Yahoo)
Medical AI tools (e.g., OpenEvidence)
Other online medical resources (e.g., UpToDate, Medscape)
Other (Specify): ______________
50. How much do you *trust* each of the following sources of information about vaccines?

a. 1 (not at all)
b. 2
c. 3 (somewhat)
d. 4
e. 5 (completely)
f. I’m not familiar with this source

News media
Social media
Web searches (e.g., Google, Bing, Yahoo)
OpenEvidence
UpToDate
Medscape
Academic/medical journals
Academic/medical institutions
Professional medical organizations
Other healthcare providers
Centers for Disease Control and Prevention (CDC)
Food and Drug Administration (FDA)
State and local public health departments
Vaccine-focused non-profit organizations
American Public Health Association (APHA)
Johns Hopkins Institute for Vaccine Safety (IVS)
National Vaccine Information Center (NVIC)
U.S. Preventive Services Task Force (USPSTF)
Vaccine Integrity Project
Other (Specify): ______________
51. How much do you trust the *vaccine recommendations* from the following sources?

a. 1 (not at all)
b. 2
c. 3 (somewhat)
d. 4
e. 5 (completely)
f. I’m not familiar with this organization’s vaccine recommendations

Advisory Committee on Immunization Practices (ACIP)
Centers for Disease Control and Prevention (CDC)
Food and Drug Administration (FDA)
Department of Health & Human Services (HHS)
My state health department
My local health department
American Academy of Pediatrics (AAP)
American Academy of Family Physicians (AAFP)
American College of Obstetricians and Gynecologists (ACOG)
American College of Physicians (ACP)
American Medical Association (AMA)
Infectious Diseases Society of America (IDSA)
Northeast Public Health Collaborative
West Coast Health Alliance
Other (Specify): ______________
52. [PN: Skip if Q50 _11 ("Centers for Disease Control and Prevention (CDC)") = f ("I’m not familiar with this source")] Please rate on scale from strongly agree to strongly disagree how you currently perceive the Centers for Disease Control and Prevention (CDC).
[PN: Randomly loop attributes listed below for rating]

a. Strongly Agree
b. Agree
c. Disagree
d. Strongly Disagree

The CDC does everything they should to protect the health of the population
The CDC is partly responsible for the illegal drug problems in this country
The CDC uses resources well
The CDC wastes money on health problems
The CDC keeps trying the same things to help the public, even when they don’t work very well
The CDC comes up with new ideas to solve health problems
The CDC bases recommendations on the best available science
The CDC is not always able to help the health of the public
The CDC is more concerned about some racial and ethnic groups than other groups
The CDC is concerned about all people, without caring about who has more or less money
The CDC accurately informs the public of both health risks and benefits of medicines
The CDC quickly helps the public with health problems
The CDC believes in what they recommend for the public
The CDC recommends things for the public that are not helpful
53. Are you familiar with the [PN: Pipe “Immunization Adverse Reaction Monitoring (IARM) System” or “Vaccine Adverse Event Reporting System (VAERS)” Assign Randomly based on emptiest bucket.] run by the CDC and FDA?

a. Yes
b. No [PN: Skip next question]
54. Have you ever made a report to the [PN: Pipe “Immunization Adverse Reaction Monitoring (IARM) System” or “Vaccine Adverse Event Reporting System (VAERS)” Assign Randomly based on emptiest bucket. Pipe-in should be consistent with the one assigned at Q53]?

a. Yes
b. No
55. Do you have everything you need to share vaccine information with your patients?

a. Yes

1) Please elaborate on why you feel this way ___
b. No

1) Please elaborate on why you feel this way ___
56. How much do you think each of the following resources would improve your vaccine-related experiences with patients?

a. 1 (not at all)
b. 2
c. 3 (somewhat)
d. 4
e. 5 (very much)

For Clinicians
An *in-person* training on evidence-based strategies for vaccine discussions with patients (with CME credit)
An *online* training module on evidence-based strategies for vaccine discussions with patients (with CME credit)
A guide summarizing scientific literature on vaccine safety concerns
Guidance on billing and reimbursement for vaccine counseling
Legal advice on vaccine-related liability
For Patients
An educational website with personalized videos about vaccines
A chatbot that answers vaccine questions
Printable handouts on vaccines
Other (Specify): ______________
57. Please share any additional opinions or thoughts that may help us provide helpful vaccine information to clinicians and patients: __________________

a. I don’t have any additional opinions or thoughts to share on this [PN: Exclusive Checkbox]

### Section Five: Additional Demographic Questions

S6. [PN: If S1 = a THEN auto punch S6=d and skip] What is your highest completed clinical degree?

a. Associate degree (for example: AA, AS)
b. Bachelor’s degree (for example: BA, BS)
c. Master’s degree (for example: MA, MS, MEng, MEd, MSW, MBA)
d. Doctorate level (for example: MD, DDS, DVM, LLB, JD, PhD, EdD)
e. Other (Specify): ______________
S7. What year did you graduate from the program of your highest clinical degree? ___
S8. Please select your race/ethnicity.

a. American Indian or Alaska Native
b. Asian
c. Black or African American
d. Hispanic or Latino
e. Native Hawaiian or Other Pacific Islander
f. White
g. Other (Specify): ______________
h. More than one of the above
i. Prefer not to answer
S9. Please select your gender identity.

a. Female
b. Male
c. Non-binary/non-conforming
d. Prefer not to answer
S10. Please select the candidate you supported in the last Presidential election.

a. Kamala Harris (Democratic Party)
b. Donald Trump (Republican Party)
c. Jill Stein (Green Party)
d. Chase Oliver (Libertarian Party)
e. None of the above
f. Prefer not to answer
S11. Would you be willing for us to contact you to schedule a virtual interview so that we can learn more about how changes made to the CDC’s immunization schedule over the past year have affected your practice and experience with patients?

a. Yes
b. No

